# Rare Comorbidity between Inflammatory Bowel Disease and Primary Biliary Cholangitis: Evidence from Causality, Shared Genetic Architecture and Transcriptomics

**DOI:** 10.1101/2023.03.01.23286611

**Authors:** Wentao Huang, Rui Jiang, Ruijie Zeng, Yuying Ma, Lijun Zhang, Shuangshuang Tong, Yanlin Lyu, Jiaxuan Wang, Felix W Leung, Weihong Sha, Hao Chen

**Affiliations:** Department of Gastroenterology, Guangdong Provincial People’s Hospital (Guangdong Academy of Medical Sciences), Southern Medical University, Guangzhou 510080, China; The Second School of Clinical Medicine, Southern Medical University, Guangzhou 510006, China; School of Medicine, South China University of Technology, Guangzhou 510006, China; Shantou University Medical College, Shantou 515041, China; Faculty of Synthetic Biology, Shenzhen Institute of Advanced Technology, Chinese Academy of Sciences, Shenzhen, China; Sepulveda Ambulatory Care Center, VA Greater Los Angeles Healthcare System, Los Angeles 91343, California, USA; University of California Los Angeles David Geffen School of Medicine, Los Angeles 90095, California, USA

**Keywords:** Inflammatory Bowel Diseases, Primary Biliary Cholangitis, Causality, Shared Genetic Architecture, Comorbidity

## Abstract

**Background:** Clinical studies have found comorbidity between Inflammatory Bowel Disease (IBD) and primary sclerosing cholangitis (PSC). Primary biliary cholangitis (PBC) is another autoimmune liver disease but the coexistence of IBD and PBC is rare. Whether there exists comorbidity between IBD and PBC and potential mechanism remains unclear.

**Methods:** We assessed the casual effect between PBC and IBD, i.e., Crohn Disease (CD) and Ulcerative Colitis (UC) independently based on genome-wide association studies (GWAS) summary statistics. By leveraging data from GWAS data, Bulk tissue RNA sequencing (bulk RNA-seq) data, and Single-cell RNA sequencing (scRNA-seq) dataset, we investigated the shared genetic architecture between IBDs and PBC. The transcriptomic expressions of shared genes were explored in patients with IBD (intestinal biopsies) and PBC (peripheral CD4^+^ T cells).

**Result:** We found a bidirectional causal relationship for PBC and IBDs using Mendelian randomization. The IBDs had been considered as the protective factors on PBC (0.87[95% confidence interval (CI): 0.81-0.93], *P* = 8.72e-5, vice versa (0.91[95% CI: 0.81-0.93], *P* = 2.65e-09). We find a consistent negative genetic correlation between PBC and IBD (LDSC: *r*_g_ = -0.2245, *P* = 2.89e-5). Cross-trait analysis yielded 9 shared risk SNPs and 7 nearest genes. In transcriptome analysis, we observed significant (*P* < 0.05) differences expression in intestinal biopsies (*PGAP3* and *DENND1B*) and in peripheral CD4^+^ T cells (*PTPN11* and *PNMT*). We identified shared tissue-specific heritability enrichment for PBC and IBD (including CD not UC) in lung, spleen and cells EBV-transformed lymphocytes and identified shared cell type-level enrichment for IBD, CD and PBC in type 1 dendritic cells, natural killer cells, CD8^+^ cytotoxic T lymphocytes in lung and activated CD8^+^ T cell in spleen.

**Conclusion:** Our study indicates that IBD and PBC are protective factors for each other and shared genetic architecture may contribute to the negative genetic correlation. These findings may explain the rare comorbidity between IBD and PBC.

## Introduction

Inflammatory bowel disease (IBD) is a group of the chronic immune-mediated idiopathic inflammation of the gastrointestinal tract and has two forms including Crohn’s Disease (CD) and Ulcerative Colitis (UC)^1–4^. IBD always experience various hepatobiliary disorders, including autoimmune liver diseases^5^. As one of autoimmune liver diseases, 70%-80% patients with primary sclerosing cholangitis (PSC) would have IBD and share a similar or common pathogenesis have been hypothesized^6^. Primary biliary cholangitis (PBC) is another autoimmune liver disease and causes chronic and persistent cholestasis in the liver, eventually resulting in cholangitis and hepatic failure without appropriate treatment^7,8^. PBC was always accompanying with various extrahepatic autoimmune diseases^9^. However, unlike PSC, PBC has little associated with IBD^5^ and there are few reports on the coexistence of PBC and IBD^10^. Patients with PBC were diagnosed after IBD had been diagnosed^11^ and coexistence with IBD had no impact on PBC outcome^12^. In addition, during rare comorbidities reports, PBC have higher rates of coexistence with CD than UC^13–15^. Whether there existed the causality and absence of comorbidities between PBC and CD or UC remain unknown.

Prior studies have revealed that genetic risk factors lead to PBC^16^ and IBD^17,18^. Genome-wide association study (GWAS) analysis had revealed strong genetic effect in the major histocompatibility complex (MHC)^19,20^ and non-MHC^19,21–23^ both PBC and IBD. These loci may contribute to the abnormal immune activation^24^ for PBC and was associated with IBD^19^. Futher investigation identified tissue-specific and cell-type-specific enrichment for IBD in CD4^+^ T cells in lung and CD8^+^ cytotoxic T cells^25,26^,which had a significant role in IBD and PBC pathogenesis^27^ including inflammation^28–30^ and immune activation^31,32^. These findings suggest that genetic factors can influence the rare coexistence of PBC and IBD but the SNP heritability enrichment for PBC remains unclear. Thus, further evaluating the shared genetic architecture may contribute to understand the potential mechanism of the rare comorbidity and explore new therapeutic targets for a precision medicine.

In the current study, based on Single-cell RNA sequencing (scRNA-seq) dataset, GWAS dataset and Genotype-Tissue Expression (GTEx) dataset, five studies were performed to dissect: (1) causal relationship between IBD and PBC relied on five Mendelian randomization methods; (2) genetic correlation between IBD and PBC; (3) shared SNP combining Heritability Estimation from Summary Statistics(ρ-HESS) and cross-trait GWAS meta-analysis; (4) transcriptomic evaluation of the risk genes; and (5) SNP heritability enrichment in the specific tissue and cell-type.

## Methods

### Data samples

#### GWAS summary statistics

GWAS summary results for IBD conclude three traits (IBD,CD and UC)^33^. GWAS summary statistics for IBD include 34,915 controls and 25,042 cases of European ancestry. The data contain 40,266 Europeans participant including 28,072 controls and12,194 cases for CD and 45,975 Europeans Europeans participant including 33,609 controls and 12,366 cases for UC. GWAS summary statistics for PBC download from the public GWAS summary statistics, which contained 8,021 European ancestry cases and 16,489 European ancestry controls^34^.The datasets used for replication are another public PBC GWAS summary statics from the EBI database (including 2,764 cases and 10,475 controls^35^). GWAS summary statistics were handled with the collection of samples and the quality control procedures, which was descripted in the publication^26,36^.

#### Bulk-tissue RNA sequencing gene expression data

The bulk-tissue RNA-seq gene expression data was available at GTEx project, contains gene expression of 53 non-diseased human primary tissues^37^, to perform the subsequent linkage disequilibrium score regression specifically expressed genes (LDSC-SEG). We chose the GTEx v6p dataset, which has been fixed^25^. For transcriptome analysis, bulk RNA-seq for IBD, CD, UC and PBC were from the Gene Expression Omnibus (GEO, http://www.ncbi.nlm.nih.gov/geo/) to evaluate the gene expression of shared risk genes in each trait. The GSE179285 dataset^38^, which collected intestinal biopsies from 254 samples that corresponded to 168 CD patients, 55 UC patients and 31 normal subjects. The gene expression matrix of IBD was a combination of CD and UC patients (N = 223). The GSE93170 dataset contained a transcriptome-wide analysis of peripheral CD4^+^ T cells derived from 6 PBC cases and 6 control subjects^39^.

#### Single-cell RNA sequencing gene expression data

Based on evidence of enrichment in heritability of tissue-level SNPs in LDSC-SEG analysis, we matched the publicly available single-cell scRNA-seq data for four enrichment tissues including spleen (N= 94,257 immune cells)^40^, lung (N = 57,020 cells)^40^, small-intestine terminal ileum (N= 53,193 epithelial cells from mouse)^41^ and hematopoietic stem and progenitor cell (HSPC, N= 7,551 adult human blood cells)^42^. For small-intestine terminal ileum data, the gene symbols were converted from mouse to human using the “EWEC” package. In total, there are 81 cell types from four tissues were included.

### Statistical analyses

#### Mendelian Randomization Analyses

We selected five MR methods including Generalized Summary-data-based Mendelian Randomization (GSMR)^43^, weighted median^44^, MR-Egger^45^, weighted mode^46^, and inverse variance weighting (IVW)^47^ to test the causality between PBC and IBD (including UC and CD), from the R packages “TwoSample MR”^48^ and “GSMR” ^43^ (*P* < 0.05) To validate the stability of the causal relationship, we also conducted a validation study using the validation group.

We performed sensitivity analysis including Leave-one-out sensitivity test, pleiotropy test, and heterogeneity test. MR-Egger regression^49^ was used to assess whether the MR-Egger intercept was different from zero and the pleiotropy of instruments^45^. The heterogeneity was assessed by Cochran’s Q statistic^50,51^. Outlier instrument was removed by leave-one-out analysis ^52,53^ and MR-PRESSO analysis^49^. In MR-PRESSO analysis, it tests those SNPs cause the heterogeneity in the estimate of the causality and estimated the global test *P* value after excluding these SNPs. Leave-one-out analysis was utilized to test whether there existed special single variant influencing the causality, which was remained and had combined effect value. We also calculated the causal effect on two traits using GSMR method. HEIDI-outlier test would remove those SNPs that show evidence of pleiotropic effects by the heterogeneity (HEIDI-outlier test < 0.05) were removed^43^. After these methods, those SNPs which show genome-wide significance (*P* < 5 × 10^−8^) from the ‘exposure’ trait was set as the instrumental variables. The validation group also performed the sensitivity analysis.

#### Heritability and Genetic Correlation

LD score regression (LDSC)^54^ (Python 2.7) was performed to evaluate the genetic correlation for different subtypes. Those SNPs which did not match the reference panel (INFO score ≤ 0.9 or MAF ≤ 0.01) would be excluded based on the LD scores that are computed from the 1000 Genomes project phase III ^55^. Based on stratified linkage disequilibrium score regression (SLDSC) with the baseline-LD model^2^, we calculated single trait SNP heritability for IBDs and PBC. Then bivariate LDSC^56^ were conducted to evaluate genetic correlations between IBDs and PBC^57^.

We also performed Genetic covariance analyzer (GNOVA)^58^ to calculate the genetic correlation and the SNP-based heritability for IBDs and PBC. GNOVA explore genetic covariance using all genetic variants shared between two diseases. We then calculated genetic correlation based on genetic covariance and variant heritability. In addition, sample overlap correction of GWAS summary statistics were corrected.

#### Evaluating of local genetic correlations using ρ-HESS

ρ-HESS^59^ is utilized to examine local SNP-heritability and genetic correlation from GWAS summary data. To identify whether there is shared genetic correlation between IBDs and PBC at the local independent region in the genome, we calculated the local genetic correlations using ρ-HESS (Python 2.7). 1,699 regions were approximately LD-independent loci with average size of nearly 1.5Mb^60^. Finally, relied on the 1000 Genomes project as reference, we estimated genetic correlation between two subtypes and the local SNP heritability for each traits^61^.

#### Cross-trait meta-analysis

We conducted two independent Cross-trait meta-analysis of GWAS summary statistics including cross-phenotype association test (CPASSOC)^62^ and multi-trait analysis of GWAS (MTAG)^63^ to identify shared risk SNPs between PBC and each of IBD, CD and UC. Based on a strong genetic correlation between two traits, MTAG performs meta-analysis of different traits when all variants have the same genetic correlation across all traits. The results of MTAG are effective with possible sample overlap between GWAS summary statistics for traits. We assessed more than 10% of SNPs were causal for each trait by calculating the value of maxFDR. CPASSOC analysis is suitable for binary traits but needs to account for potential false positives due to sample overlap. Shared risk SNPs were determined to be significant (*P* < 5×10^−8^) in both CPASSOC and MTAG. These independent SNPs were identified by LD clumping (r^2^□< 0.001 within 10,000-kb windows) in Plink. Eventually, share risk SNP were identified significant if they also surpassed Bonferroni-correction (N= 1,562, *P* < 3.2×10^−5^) for local genetic correlation in ρ-HESS analysis.

#### Transcriptomic evaluation of the risk genes

We evaluated and compared whether the shared risk genes obtained from the above analysis exhibited significant (*P* < 0.05) differences in genes expression occurred in IBD, CD, UC, PBC patients and normal subjects. We selected the GSE179285 dataset^38^, which is a collection of 254 samples that corresponded to 168 CD patients, 55 UC patients and 31 normal subjects. The gene expression matrix of IBD was a combination of CD and UC patients (N = 223). The GSE93170 dataset contained a transcriptome-wide analysis derived from 6 PBC cases and 6 control subjects. The “ggplot2” and “ggpubr” packages were used for the estimation of differences among the gene expression profiles and visualization.

#### LD score regression applied to specifically expressed genes (LDSC-SEG)

We implemented LDSC-SEG^25^ to infer the enrichment for SNP heritability and the related tissue-specific enrichment in IBD, CD, UC and PBC, using specifically expressed genes sets in tissues and GAWS summary statistics for each trait. We utilized 1000 Genomes Project in Phase 3 as the reference panel and only HapMap 3 SNPs with minor allele frequency (MAF) > 0.05 were included. The gene sets utilized in LDSC-SEG were defined by Finucane et al. for the genotype tissue expression (GTEx) project with 53 tissue types. According to the t-statistic, the top 10% of candidate genes were selected as the set of genes specifically expressed in tissues. The regression coefficient p-values were determined from the coefficient Z-score, and the false discovery rate (FDR) <?0.05 was retained to represent the significance of enrichment tissues across PBC and each trait of IBD, CD and UC.

#### Multi-marker Analysis of Genomic Annotation (MAGMA)

Cell type-specific enrichment was performed by MAGMA method^64^, which converted SNP associations into gene-level associations and estimated enriched cell types via differential expression data from the input scRNA-Seq data. We matched the publicly available single-cell scRNA-seq data for four tissues including spleen (N= 94,257 immune cells)^40^, lung (N = 57,020 cells)^40^, small-intestine terminal ileum (N= 53,193 epithelial cells from mouse)^41^ and HSPC (N= 7,551 adult human blood cells, GSE137864)^42^ which were enriched in LDSC-SEG. MAGMA enrichment results were analyzed with “MAGMA.Celltyping” R package (version 2.0.7) and all scRNA-Seq data for tissues were processed with the “EWEC” R package (version 1.5.8)^65^. Results with FDR > 5 × 10^−3^ indicated significant and independent genetic signals relative to cell type.

## Results

### Causality between IBD and PBC

The bi-directional MR were conducted to assessed the casual association between IBD and PBC. Our results found the causal effect of IBD on PBC in five methods (0.87[95% CI: 0.81-0.93], *P* = 8.72e-5) (**Fig. 1 and Supplementary S4**). IBD played the protective factor to the PBC. No significant horizontal pleiotropy for the causality of IBD on PBC existed (MR-PRESSO global test *P* = 0.089; MR-Egger intercept = 3.56e-4, SE = 0.013, *P* = 0.98; **Supplementary Fig.S1**).

**Figure. 1.**
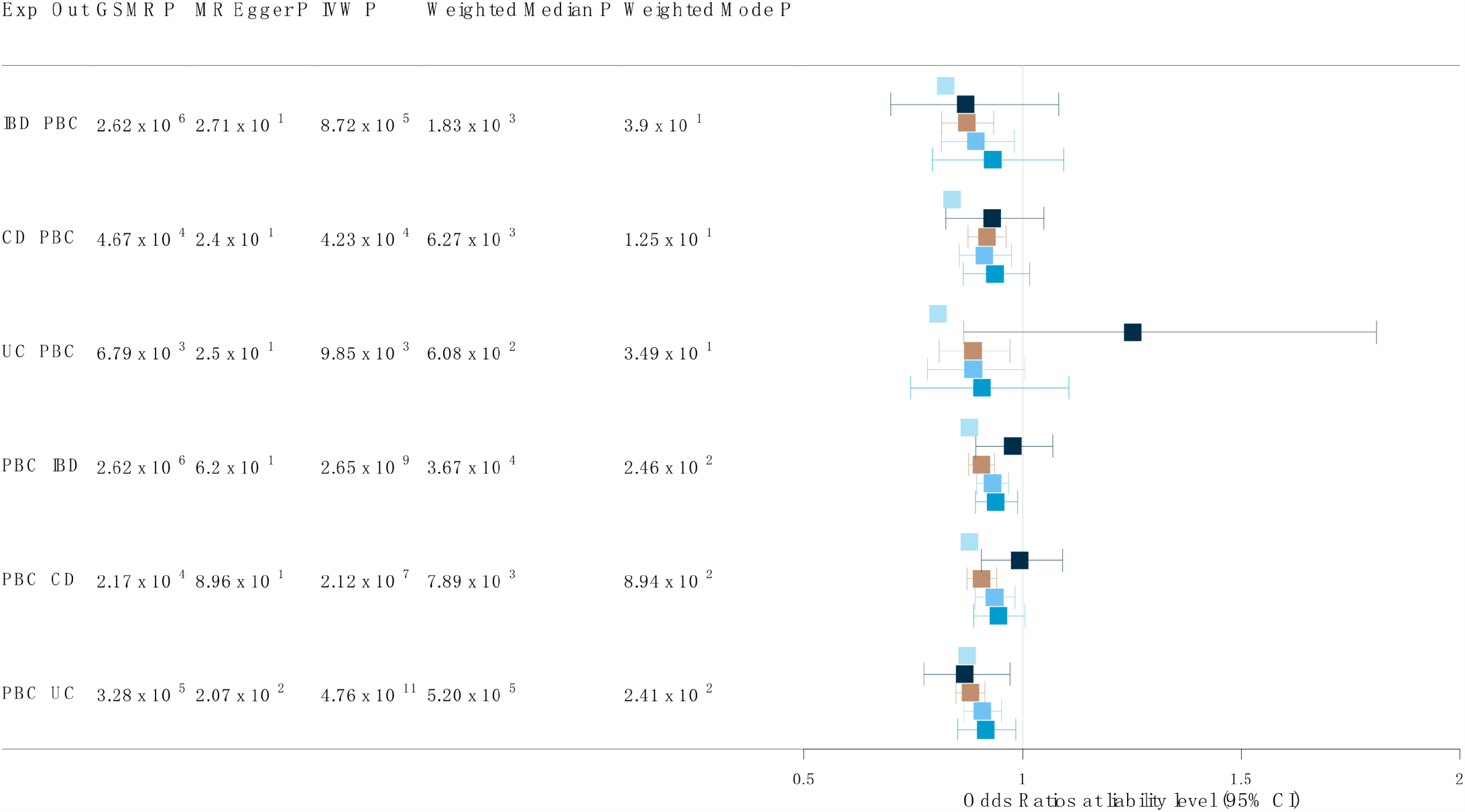
The estimation of causality between PBC and IBD (Including CD and UC) based on five Mendelian Randomization analysis methods. Five boxes represent GSMR: Generalized Summary-data-based Mendelian Randomization, MR Egger, IVW: Inverse variance weighting, Weighted Median and Weighted Mode from the top to bottom. IBD: inflammatory bowel disease, CD: Crohn’s disease, UC: ulcerative colitis, PBC: primary biliary cholangitis.

In the reverse analyses, five methods found consistent evidence for causality of PBC on IBD (**Fig.1 and Supplementary S4**). PBC was also been considered as the protective factor to the IBD (0.91[95% CI: 0.81-0.93], *P* = 2.65e-09). IBD played the protective factor to the PBC. There was no horizontal pleiotropy for the causality of IBD on PBC (MR-PRESSO global test *P* = 0.09; MR-Egger intercept = -0.0178, SE = 0.0101, *P* = 0.094; **Supplementary Fig.S5**). The causal association was verified using the validation sets (**Supplementary Fig.S8**).

### Causality between PBC and two subtypes of IBD

We further explored casual effect for two phenotypes of IBD (CD and UC) on PBC. There existed the causality of CD and UC on PBC. (CD: 0.92 [95% CI: 0.88-0.96], *P* = 4.23e-4; UC: 0.89[95% CI: 0.81-0.97], *P* = 0.00985484) (**Fig. 1 and Supplementary Fig.S4**). Both two subtypes played the protective factor to the PBC. No significant horizontal pleiotropy for the causality of CD on PBC was tested (CD: MR-PRESSO global test *P* = 0.218; MR-Egger intercept = -0.0025, SE = 0.011, *P* = 0.82; UC: MR-PRESSO global test *P* = 0.223; MR-Egger intercept =-0.052, SE = 0.02742005, *P* = 0.07; **Supplementary Fig.S2-S3**).

In the reverse analyses, five methods found consistent evidence for causal effect of PBC on CD and UC (**Fig.1 and Supplementary S4**). PBC was also been considered as the protective factor to the two subtypes (CD: 0.88[95% CI:0.85-0.91], *P* = 4.76e-11; UC: 0.88[95% CI: 0.85-0.91], *P* = 4.76e-11). CD and UC played the protective factor to the PBC. There was not significant horizontal pleiotropy for the causality of CD and UC on PBC (CD: MR-PRESSO global test *P* = 0.313; MR-Egger intercept = -0.022, SE = 0.011, *P* = 0.05; UC: MR-PRESSO global test *P* = 0.09; MR-Egger intercept =0.0033, SE = 0.012, *P* = 0.79; **Supplementary Fig.S6-S7**). The causal effect was verified using the validation sets ((**Supplementary Fig.S8**).

### Estimation of Genetic correlations between IBDs and PBC

We calculated the SNP heritability for IBDs and PBC using SLDSC^54^ with the baseline-LD model^66^. the SNP heritability for IBDs^26^ and for PBC was consistent with the previous study. The results of bivariate LDSC show that IBD including CD and UC have negative correlation with PBC (**Supplementary Table 1**). To get a robust result, we conducted GNOVA and explored a consistent result between PBC and IBDs after correcting sample overlap (**Supplementary Table 1**).

### Local genetic correlations between IBD, UC, CD and PBC

Local genetic correlation was evaluated for 1,561 genomic partitions and Bonferroni significant regions were assessed (**Supplementary Table S1-S3)**. Analysis of local genetic correlation revealed loci of significant local genetic covariance to four traits (**Supplementary Fig. S9-S11**). Polygenicity was higher for PBC than for IBDs. There was close agreement in the average local genetic correlation in regions harboring PBC-specific loci or IBDs-specific loci (**Fig. 2**). We estimated the local single-trait SNP heritability for PBC and IBDs (PBC: 0.255, IBD:0.466, CD:0.497, UC:0.296). Genome-wide local genetic correlations tested by HESS were all largely consistent with genetic correlation estimated by LDSC and GNOVA (IBD-PBC: -1.12, CD-PBC: -1, UC-PBC: -1.3) between IBDs and PBC. Polygenicity was higher for IBDs than for PBC (**Supplementary Fig.S12**). The potential correlation for sharing of genetic variation may exist in the whole genome.

**Figure. 2.**
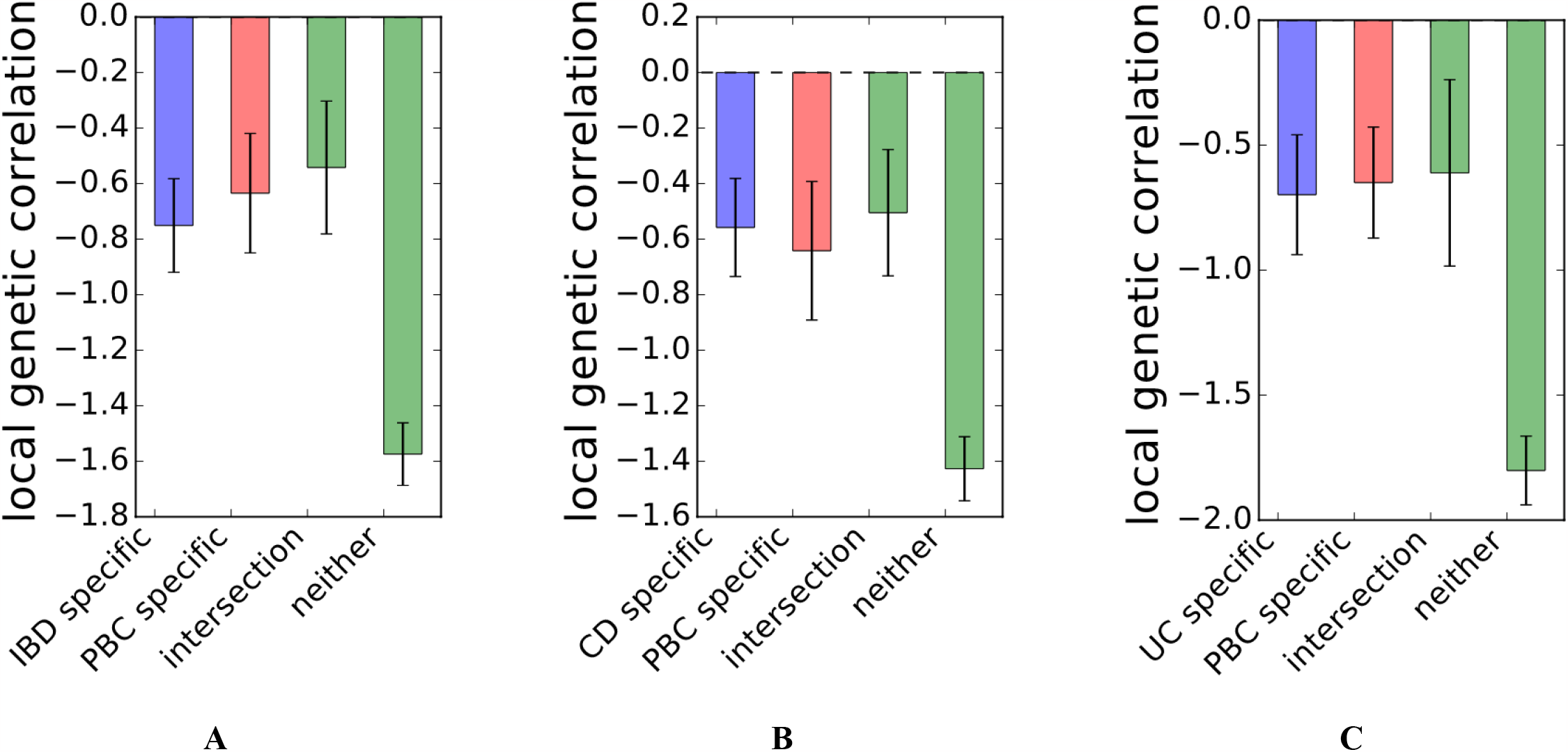
Local genetic correlation at loci ascertained for GWAS hits (p<5e-6) specific to PBC and IBD(A), CD (B) and UC (right), respectively. IBD: inflammatory bowel disease, CD: Crohn’s disease, UC: ulcerative colitis, PBC: primary biliary cholangitis.

### Shared risk SNPs were identified between PBC and each trait of IBD, CD and UC

With significant genetic correlations between PBC and each of IBD, CD and UC, we utilized two complementary cross-trait meta-analysis approaches to identify shared risk SNPs between IBDs and PBC. With significant local genetic correlation, we identified that six SNPs (rs11583319, rs7583409, rs6580223, rs11066301, rs407307, rs5757587) were significantly (*P* < 5×10^−8^) associated with the joint phenotype IBD-PBC, three SNPs (rs67236816, rs407307, rs5757587) were significant in the CD-PBC cross-trait GWAS, three SNPs (rs10752745, rs12946510, rs5757587) were significant in the UC-PBC. We totally obtained 9 candidate shared risk SNPs including two previously discovered SNPs (rs6580223 and rs12946510) associated with IBD and/or PBC and seven potential novel SNPs. The maxFDR values for MTAG analyses of PBC and each of IBD, UC, and CD were 0.014, 0.005, 0.004 and 0.026 respectively. The lists of shared risk SNPs were summarized in **Table 2**.

**Table 1.** Heritability and genetic correlation estimated for PBC and IBD including CD and UC using LDSC and GNOVA. PBC: primary biliary cholangitis, IBD: inflammatory bowel disease, UC: ulcerative colitis, CD: Crohn’s disease, LDSC: LD score regression, GNOVA: Genetic covariance analyzer.

**Table 2.** Shared risk SNPs associated with cross-trait PBC and IBD (or UC or CD) presented by MTAG, CPASSOC and ρ-HESS. PBC: primary biliary cholangitis, IBD: inflammatory bowel disease, UC: ulcerative colitis, CD: Crohn’s disease, SNP: single nucleotide polymorphisms, BP: base pair, MTAG: Multi-Trait Analysis of genome-wide association study (GWAS), CPASSOC: Cross Phenotype Association, HESS: Heritability Estimation from Summary Statistics, CHR: chromosome.

### Differential expression of the risk genes and correlation of gene expressions

We further evaluated the mRNA expressions of candidate risk genes (*DENND1B, DNMT3A, NDFIP1, PTPN11, PNMT, PGAP3*) in PBC and each of IBD, CD and UC patients. Significant (*P* < 0.05) differences of *PGAP3* expression and *DENND1B* expression were observed in patients with UC (*P*_*PGAP3*_ =0.013, **Supplementary Fig. S13C**; *P*_*DENND1B*_ = 1.5×10^−3^, **Supplementary Fig. S13D**) and control subjects. The expression of *PGAP3* was significantly lower than that of the control in IBD samples (*P* = 0.021, **Supplementary Fig. S13A**), CD patients (*P* = 0.035, **Supplementary Fig. S13B**) and UC samples (*P* = 0.013; **Supplementary Fig. S13C**). In PBC patients, the expression of *PTPN11* was significantly higher compared with the control group (*P* = 8.7×10^−3^, **Supplementary Fig. S13E**), and conversely, the expression of *PNMT* was decreased (*P* = 0.041, **Supplementary Fig. S13F**).

### SNP heritability enrichment of SNPs in IBD, CD, UC and PBC-related tissues

We used LDSC-SEG to investigate tissue-specific enrichment of SNP heritability for IBD, UC, CD and PBC. We detected a significant enrichment at an FDR > 0.05 threshold for tissues, IBD and CD exhibited FDR-significant enrichment in lung, spleen, whole blood and small intestine-terminal ileum tissues, CD also enriched in cells EBV-transformed lymphocytes. For PBC, we observed significant enrichment in three tissues including lung, spleen and cells EBV-transformed lymphocytes. Lung and spleen were commonly associated tissues with IBD, CD and PBC **(Figure 3A-D)**. However, there was no significant evidence for a specific enriched tissue for UC. The magnitude of SNP heritability enrichment in these tissues are presented in **Supplementary Table S4 and Supplementary Figure S14**.

**Figure. 3.**
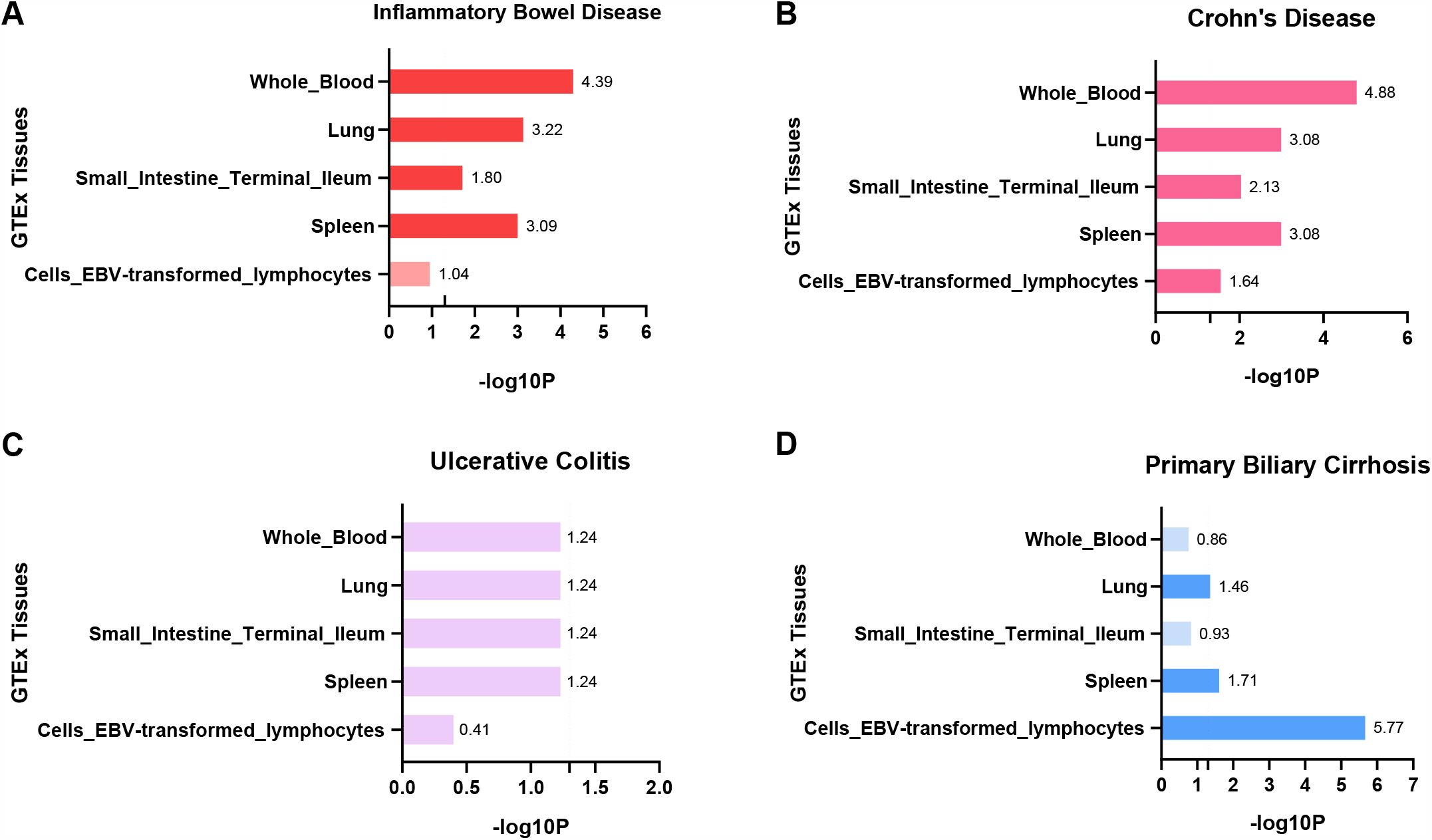
Tissue type-specific enrichment SNP heritability for (A) inflammatory bowel disease, (B) Crohn’s disease, (C) ulcerative colitis and (D) primary biliary cholangitis. The x axis represents negative log10 FDR-adjusted *P*-values for each test, with the grey dotted lines indicating a 5% FDR. FDR: false discovery rate, SNP: single nucleotide polymorphism.

**Figure. 4.**
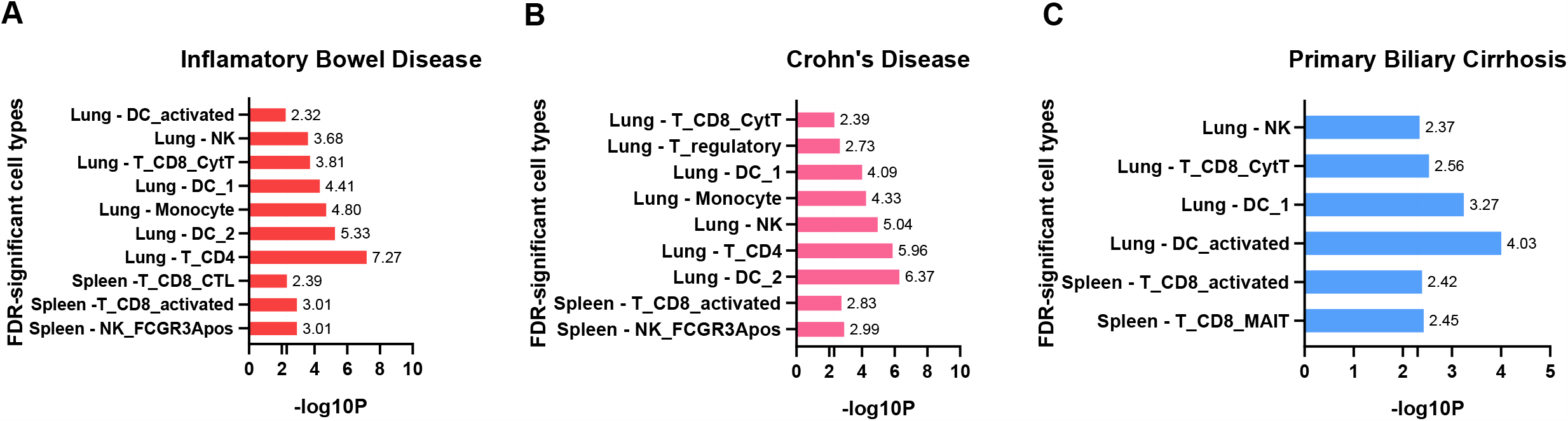
Cell type-specific enrichment for (A) inflammatory bowel disease, (B) Crohn’s disease and (C) primary biliary cholangitis, analyzed by MAGMA method. The x axis represents negative log10 FDR-adjusted *P*-values for each test, with the grey dotted lines indicating a 5% FDR. FDR: false discovery rate, SNP: single nucleotide polymorphism, MAGMA: multi-marker analysis of GenoMic annotation.

### Cell type-level enrichment in IBD, CD, UC and PBC

To identify the enrichment of cell types in spleen, lung tissues, we performed MAGMA to test for a correlation between cell type expression specificity and GWAS summary statistics of IBD, CD, UC and PBC. Of the enrich tissues identified in previous LDSC-SEG for IBD, CD, UC and PBC. We detected an enrichment at an FDR > 5 × 10^−3^ threshold for enrichment tissues. In lung tissues, we observed enrichment in CD4^+^ T cells, type 2 dendritic cells (DC2), Monocyte cells, Nature killer (NK) cells, CD8^+^ cytotoxic T lymphocytes for IBD; and DC2, CD4^+^ T cells, NK cells, Monocyte cells, dendritic cells (DC1) DC1, regulatory T cells, CD8^+^ cytotoxic T lymphocytes for CD; and activated DC cells, DC1, CD8^+^ cytotoxic T lymphocytes, NK cells for PBC (**Supplementary Fig. S15**). In spleen tissues, NK_FCGR3Apos, activated CD8^+^ T cells, CD8 cytotoxic T lymphocytes were enriched in IBD; NK_FCGR3Apos and activated CD8^+^ T cells were enriched in CD; CD 8 mucosa-associated invariant T cell, activated CD8^+^ T cells were enriched in PBC (**Supplementary Fig. S16**). We observed shared cell type-level enrichment for IBD, CD and PBC in DC1, NK cells, CD8 cytotoxic T lymphocytes in lung and activated CD8^+^ T cell s in spleen **(Supplementary Fig. 4A-4C)**. None of the cell types in small-intestine terminal ileum and HSPC showed significant specificity in all four phenotypes. The results of cell-specific types show enrichment were described in detail in **Supplementary Table S5 and Supplementary Figure S15-S18**.

## Discussion

To our knowledge, we firstly confirmed mutual protective association and negative genetic correlation between IBD and PBC. Combining novel GWAS datasets, tissue and cell-type specific expression data, we explored the shared genetic architecture between IBD and PBC to explain the rare comorbidity for two diseases.

In the previous study, the coexistence of both conditions is rare. A retrospective analysis (2006-2016), the largest series reported to date of PBC-associated IBD, reported six patients with PBC-associated IBD^11^. The number of cases reported by other clinicians is similar^67–70^. During the current study, based on the latest GWAS summary data, we firstly performed Mendelian randomization studies and identified a consistent bidirectional causality between IBD and PBC. The mutual protective roles may explain the rare comorbidity between IBD and PBC. These results provide the evidence that the genetic factors may contribute to the occurrence and development of comorbidity.

We identified the genetic correlation to provide evidence for the clinical arrange, drug discovery and disease prediction of comorbidity. The previous study found that the chromosome loss played an important role in the occurrence and the complication of PBC^71,72^. The previous GWAS study had identified chromosome 11q23.1^73^ and 3q13.33^74^ as susceptibility locus. These results may indicate that pleiotropic effects on the these traits exist in numerous genetic variants across the genome for a consistent *r*_g_ estimated by ρ-HESS, LDSC and GNOVA^26^.

To explore the shared risk SNPs between PBC and each of IBD, CD and UC, we combined HESS with joint analysis of GWAS summary results from related traits. We found two previously reported SNPs and seven novel SNPs with biologically plausible genetic associations with IBD-PBC, CD-PBC or UC-PBC. CD4^+^ T cells play a significant role in immunological dysfunction observed in PBC^75^ and IBDs^76^. The expression of nearest genes to the loci in CD4^+^ T cells were associated with the inflammatory responses which may mediate the protective mechanism between PBC and IBDs. Polymorphisms in *NDFIP1* are associated with the development of IBD^77^ and the critical function for *NDFIP1* in peripheral tolerance is as an induced, cell-autonomous brake against effector CD4^+^ cells differentiation^78^. The expression of *PGAP3* reduced in noninflamed ileum biopsies from individuals in the overall IBD cohort^79^. The expression of *PGAP3* in CD4^+^ cells of PBC is reduced at the transcriptome level. GWAS analysis found that noncoding polymorphisms in the de novo methyltransferase enzyme *DNMT3A* locus are associated with a higher risk for IBD^80^. The suppression of *DNMT3A* mRNA levels^81^ have an effect on the genetic risk for IBD. Our results proved this point in transcriptome validation. The prevous study had indicated that the expression levels of *PTPN22, PTPN11*, and *PTPN2* play a significant role in actively inflamed intestinal tissue. Loss of *PTPN11* is also associated with increased colitis severity in vivo^82^. The downregulation of *PTPN11* would reduce the production of reactive oxygen species and protect intestine against oxidative stress-induced injury^83^. In PBC patients, the expression of *PTPN11* is also reduced at the transcriptome level. Therefore, the expression of the nearest gene to the risk SNP in CD4^+^ T cells may contribute to the protective mechanism between PBC and IBDs.

In the other hand, the functional variant drives the nearest gene expression. Hitomi et al identified the rs12946510 affects the pathogenesis of PBC by altering *FOXO1* binding affinity in vitro to affect functional variation in gene expression^84^. Meanwhile, *FOXO1* plays a role in mucosal barrier injury of inflammatory bowel disease through TLR4/MyD88/MD2-NF-κB inflammatory signaling^85^. And down-regulation of *FOXO1* leads to over-production of Th17 cells and increased inflammatory responses^86^. *DENND1B* has been previously reported as one of the susceptibility gene regions that are associated with IBD(rs12740041)^87^ especially CD and PBC((rs2488393))^88^. However, our results found that *DENND1B* expression was significantly reduced in UC at the transcriptomic level. Because we identified a novel risk SNP (rs11583319) to *DENND1B and this may* explain the difference. Four SNPs (rs67236816, rs407307, rs5757587, rs10752745) and three genes (*PNMT, PGAP3* and *LOC100996583*) that have never been reported to be genetically associated with PBC or IBD, including CD and UC. In our analysis, *PNMT* are notable for its depressed expression in PBC and elevated expression in IBD including UC and CD. These results indicated that the biological mechanisms of these novel SNPs and genes underlie disease susceptibility to PBC and each trait of IBD, CD and UC are required to be investigated by further functional research.

Although IBD and PBC primarily affect the small intestine and liver, respectively, there is growing evidence that other tissues and organs might also contribute to their pathogenesis. Tissue enrichment analysis illustrated that IBD, CD and PBC were mainly enriched in immune tissues such as lung, spleen and EBV-transformed lymphocytes. Lung and spleen presented co-enrichment in IBD, CD and PBC, but not UC, suggesting dissimilar shared etiologies between PBC-UC and PBC-CD in different tissues. IBD, including CD and UC, is characterized by chronic inflammatory conditions in the intestinal tract, mainly resulted from a dysfunctional regulation of the immune system and an imbalance of the intestinal microbiota. Likewise, PBC is also characterized by the immune-mediated progressive cholestatic liver disease^89^. It has been reported that the possible immune pathways of IBD leading to lung lesions include dysfunctional immune cell homing, systemic inflammation, and microbial dysbiosis^90^. Changes in spleen size in patients with PBC may be useful to predict advanced fibrosis^91^. Previous studies have often examined tissue in the small intestine of IBD patients or liver of PBC patients. Our results suggest that IBD-PBC and CD-PBC might have a common pathogenic mechanism in common enriched tissues, such as lung and spleen. Although SNP enrichment for UC in tissue special levels was not significant, based on the strong genetic correlation and the risk SNP was identified, we suspected UC might have shared genetic overlaps with PBC, which deserves further study.

We then extended MAGMA to the single cell-level enrichment and identified three cell types (DC1, CD8^+^ cytotoxic T lymphocytes, NK cells) in lung and activated CD8^+^ T cell in spleen with significant heritability enrichments in IBD-PBC and CD-PBC. Inflammatory products induce DC cell maturation, and DC activation is a contributing factor to the development of IBD^92,93^. DC cells can induce a large number of effector T cells, and CD8^+^ T cells are involved in the pathogenesis of IBD^94^. The decrease of CD8^+^ T cell responses to commensal bacteria leading to Trm deficiency in the colon could lead to IBD^95^. Consistent with previous research results, DC cells, T cells and NK cells all played a role in the pathogenesis of PBC^96^. Activated T cells, particularly CD8^+^ T cells, play a critical role in the destruction of biliary cells and mediate the pathogenesis of PBC in both human and murine models^97^.There existed no difference for the numbers of DC(including DC1 and DC2) and the expression of human leukocyte antigen DR (HLA DR) among PBC patients and controls^98^. NK cell-mediated innate immune responses may be crucial in the initial stages of PBC. An analysis of NK cells isolated from the spleen of PBC patients suggested that NK cells might activate autoreactive CD4^+^ T cells by secreting IFN-γ to destroy biliary epithelial cells^96^. CD4^+^ T cells were enriched at IBD including CD. Certainly, as mentioned in the preceding text, CD4^+^ T cells mediated the protective mechanism through the risk SNP. These results may provide evidence that the difference in immune activation may also contribute to explain the protective mechanism.

There are some limitations in our study. Firstly, to prevent population stratification, all datasets are only from European population. Hence, there existed limited generalizability to these other populations. Secondly, although we revealed the potential shared genetic architecture but how the shared biological pathways work warrants further research. Thirdly, the sample size for PBC was relatively smaller, which might influence the results to some extent.

## Conclusion

In summary, we firstly confirmed the mutual protective correlation and negative genetic correlation between IBD and PBC. We identified novel shared risk SNPs and further explored the shared genetic overlap in tissue and cell-type levels between IBD and PBC. These findings may help to better explain the reason of rare comorbidity between IBD and PBC and may contribute to the targeted treatment and clinical management.

## Supporting information

Supplemental Table S1. Summary of local genetic correlations between PBC and each of IBD, CD, and UC

Supplemental Table S2. Summary of local genetic correlations between PBC and CD

Supplemental Table S3. Summary of local genetic correlations between PBC and UC

Supplementary Table S4. SNP heritability enrichment in 53 GTEx tissues for PBC and IBD, including CD and UC.

Supplementary Table S5. Summary of cell-types enrichment analysis for PBC, IBD, including UC and CD, conducted by the MAGMA.

## Data Availability

All data produced are available online at

## Data source

GWAS summary statistics for PBC are available by application from: https://www.ebi.ac.uk/gwas/.

GWAS summary statistics for PBC for replication are available by application from: https://gwas.mrcieu.ac.uk/.

GWAS summary statistics for IBD are available by application from: https://www.ebi.ac.uk/gwas/.

GEO dataset:

PBC: https://www.ncbi.nlm.nih.gov/geo/query/acc.cgi

IBD: https://www.ncbi.nlm.nih.gov/geo/query/acc.cgi?acc=GSE179285

GTEx-seq: https://alkesgroup.broadinstitute.org/LDSCORE/LDSC_SEG_ldscores

scRNA-seq data:

Whole blood:

Lung and Spleen: https://doi.org/10.1186/s13059-019-1906-x;

Small Intestinal Epithelium:

https://singlecell.broadinstitute.org/single_cell/study/SCP44/small-intestinal-epithelium

## Code availability

LDSC: https://github.com/bulik/ldsc.

PLINK: https://www.cog-genomics.org/plink/1.9.

MTAG: https://github.com/JonJala/mtag.

CPASSOC: http://hal.case.edu/~xxz10/zhuweb/.

GSMR: http://cnsgenomics.com/software/gsmr/.

TwoSampleMR:https://mrcieu.github.io/TwoSampleMR/.

LDSC-SEG: https://github.com/bulik/ldsc/wiki/Cell-type-specific-analyses

MAGMA Celltyping: https://neurogenomics.github.io/MAGMA_Celltyping

## Author contributions

WTH, RJ, RJZ and HC designed, revised and supervised the study; WTH, RJ, RJZ and XJW analyzed and organized the data; YYM, LJZ, SST and YLL generated the figures and tables; WTH, RJ, RJZ and HC wrote this manuscript; All authors reviewed and approved the final manuscript.

## Acknowledgements

We gratefully acknowledge the contributions from public available databases and the participants who contributed to those studies.

