## Supplementary Figure for "Rare Comorbidity between Inflammatory Bowel Disease and Primary Biliary Cholangitis: Evidence from Causality, Shared Genetic Architecture and Transcriptomics"

### Slide 1
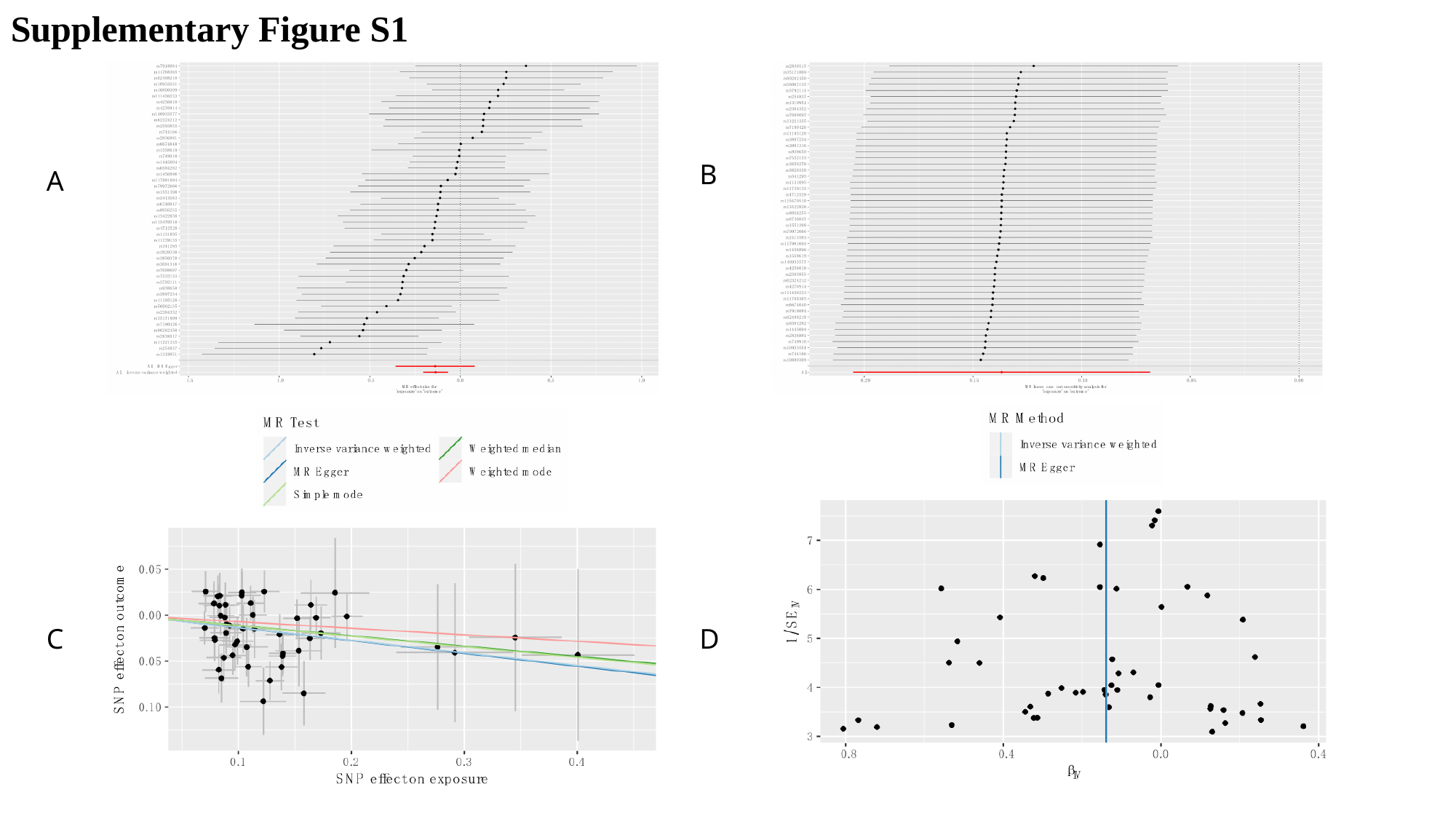

Supplementary Figure S1
B
A
C
D

### Slide 2
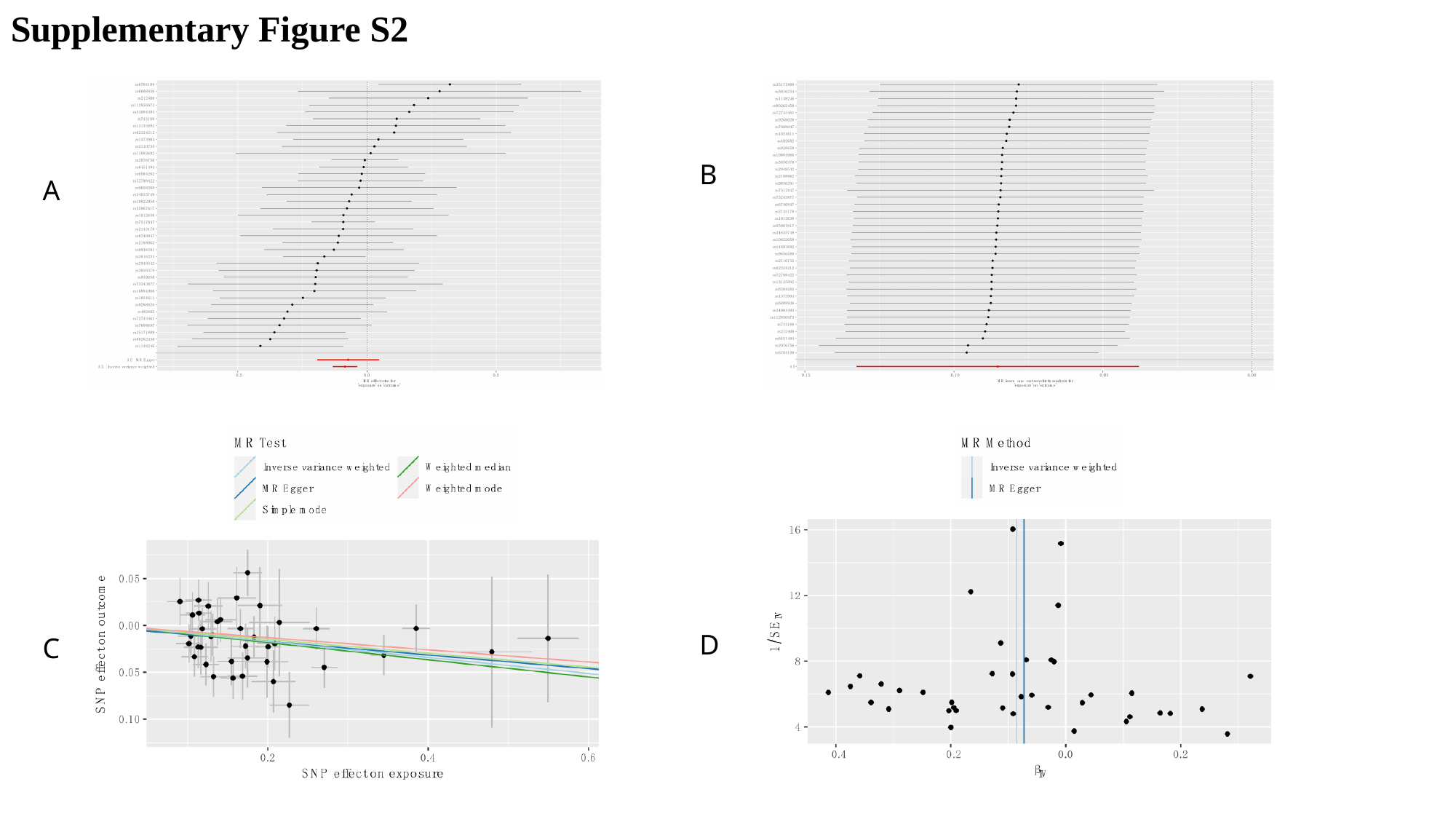

Supplementary Figure S2
B
A
D
C

### Slide 3
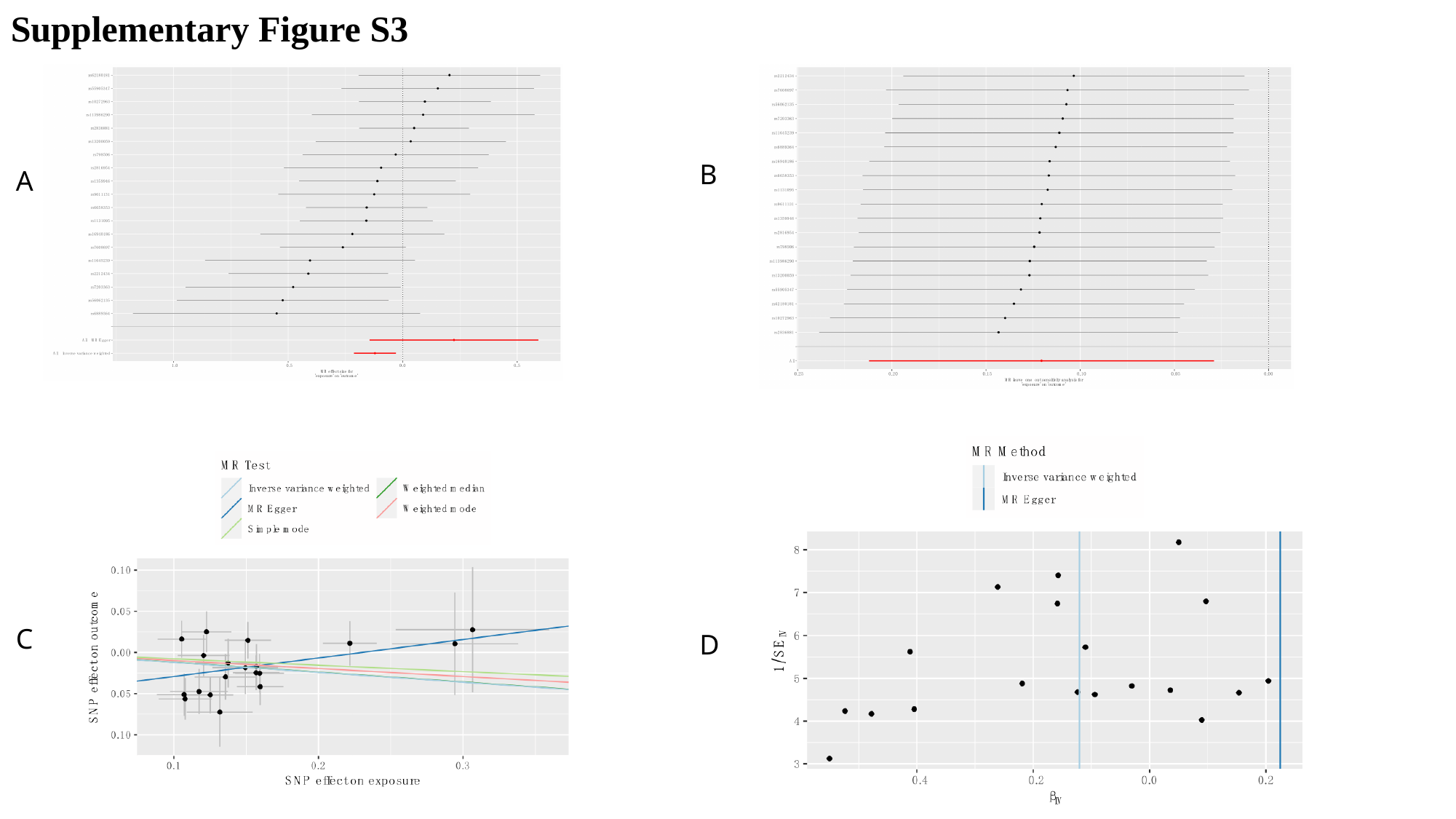

Supplementary Figure S3
B
A
C
D

### Slide 4
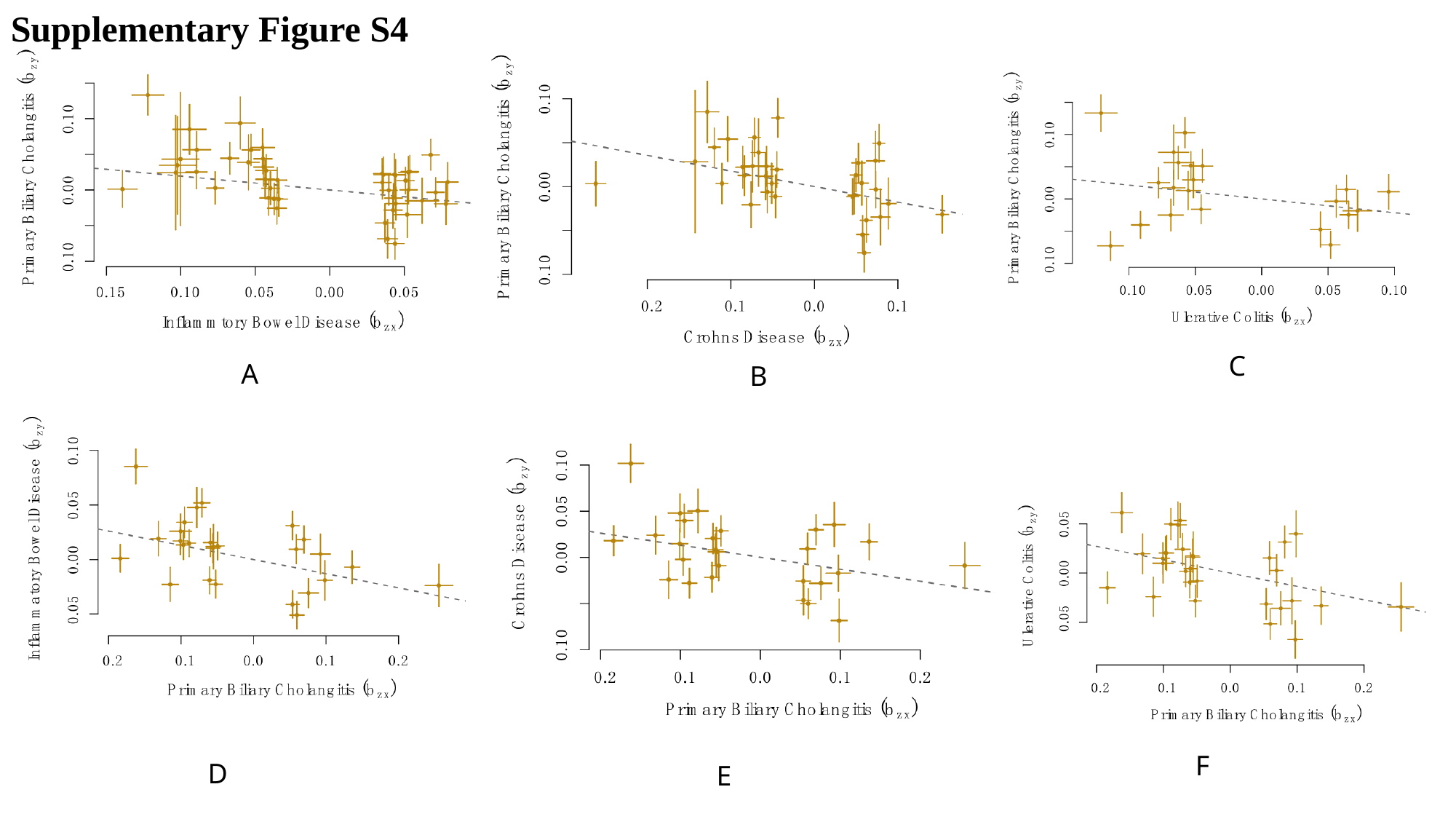

Supplementary Figure S4
C
A
B
F
D
E

### Slide 5
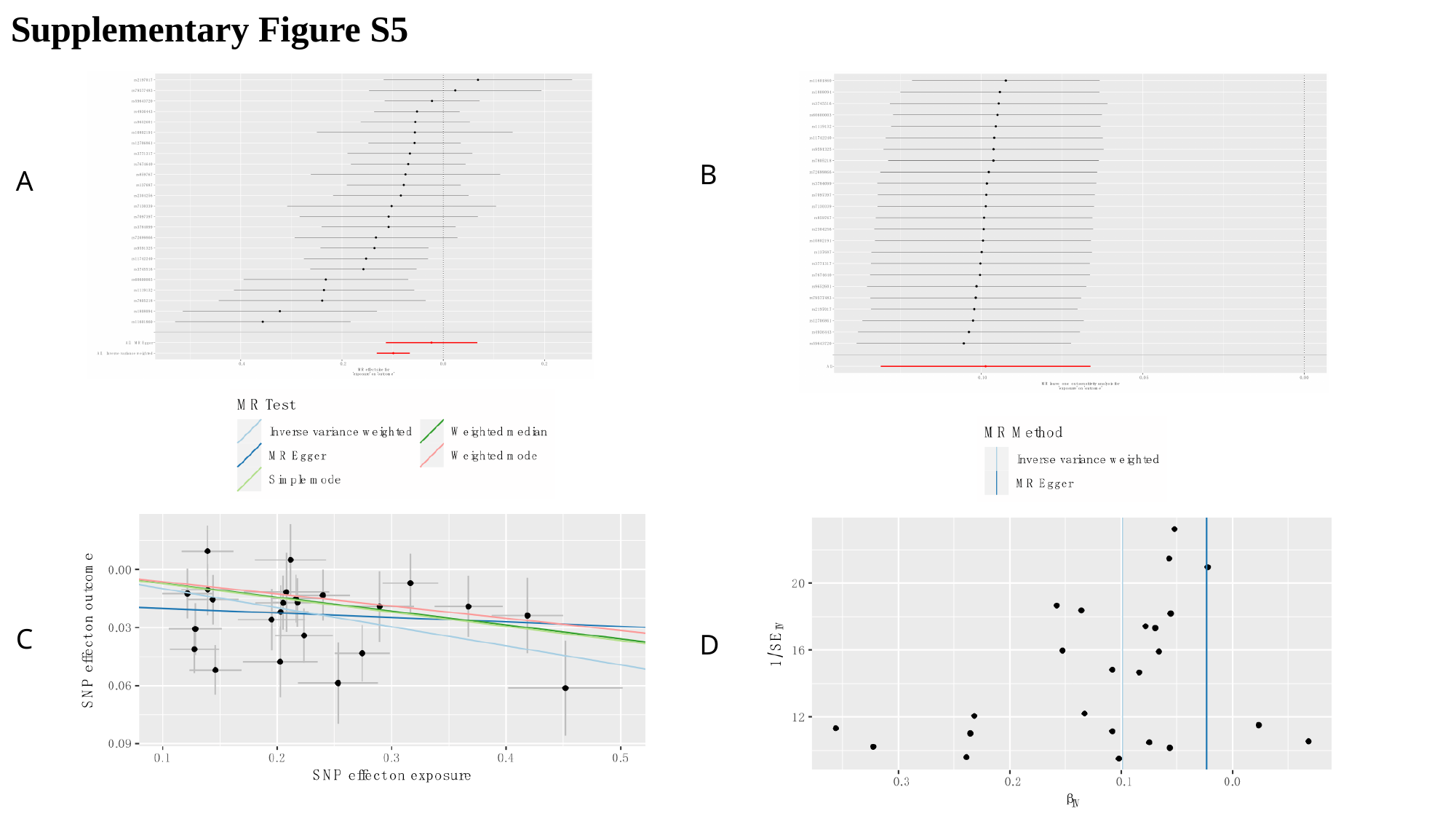

Supplementary Figure S5
B
A
C
D

### Slide 6
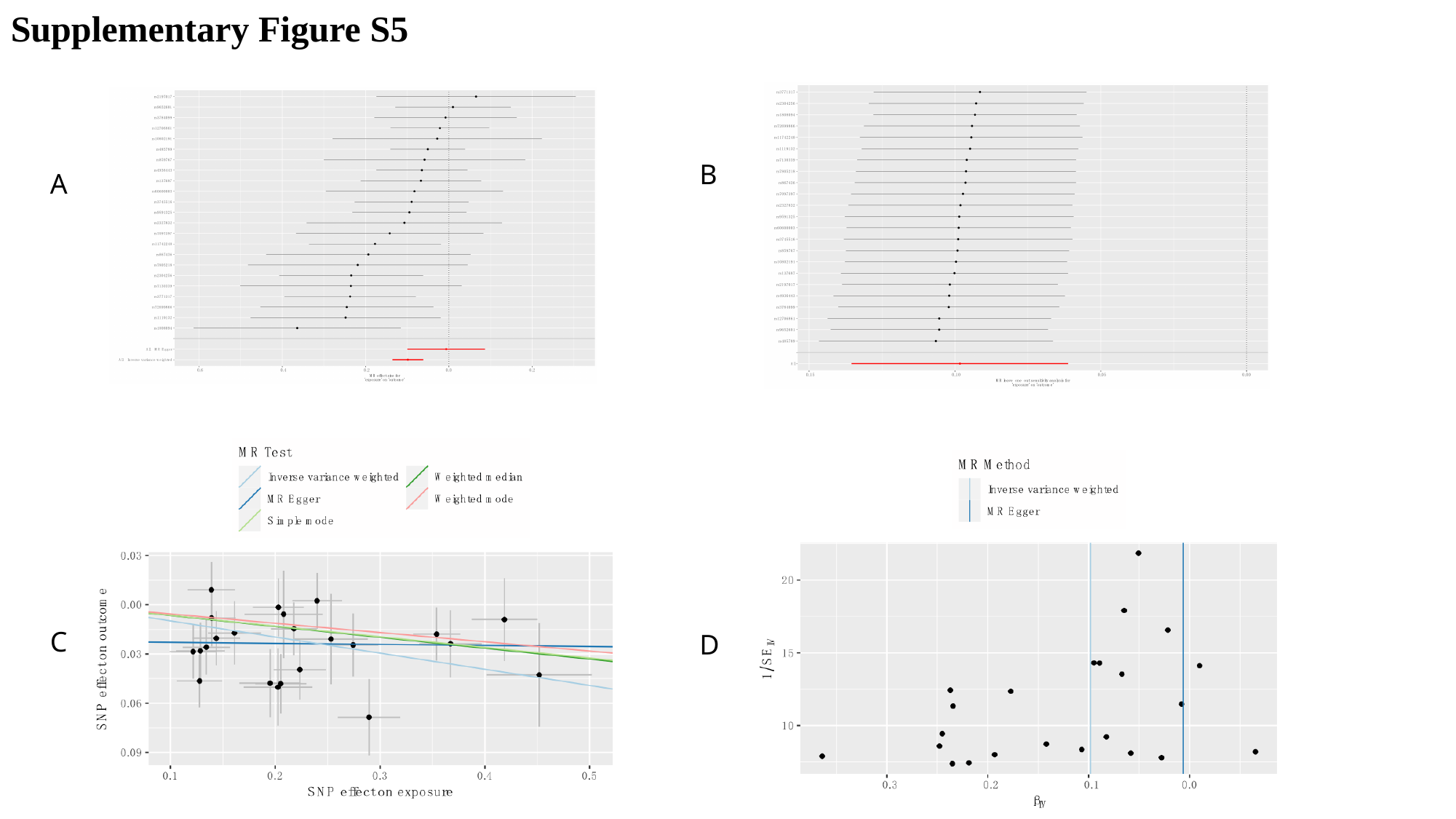

Supplementary Figure S5
B
A
C
D

### Slide 7
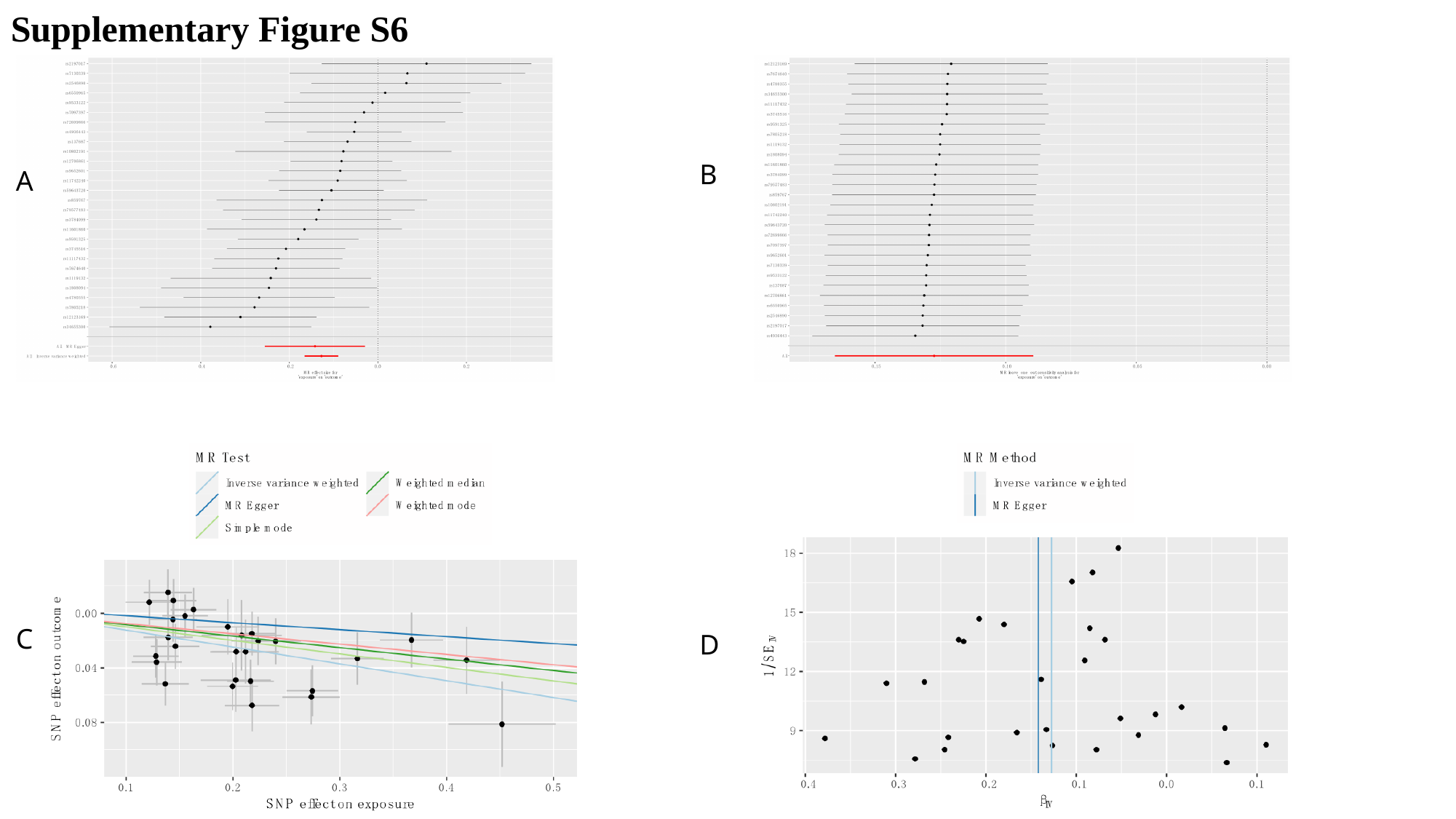

Supplementary Figure S6
B
A
C
D

### Slide 8
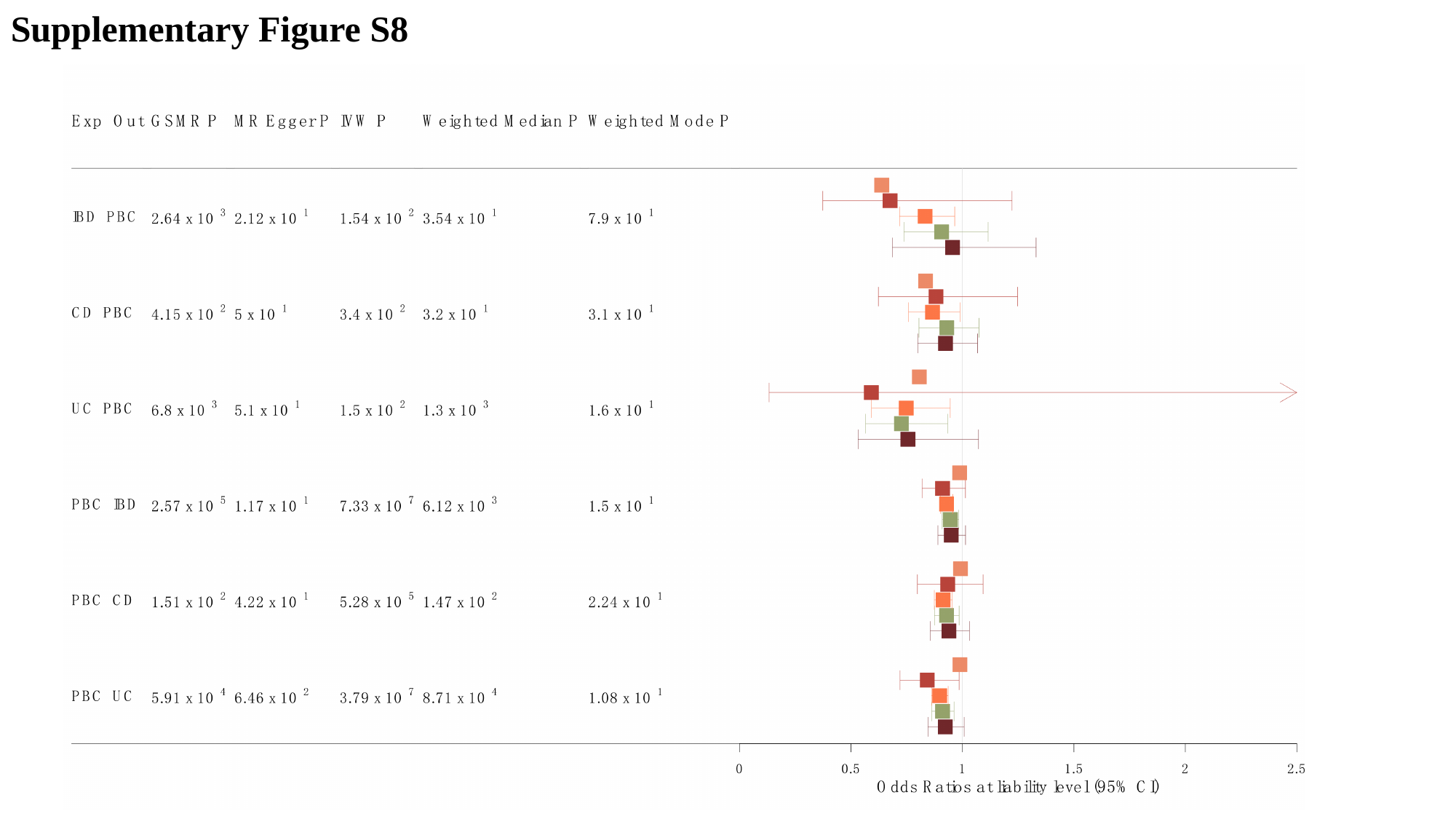

Supplementary Figure S8

### Slide 9
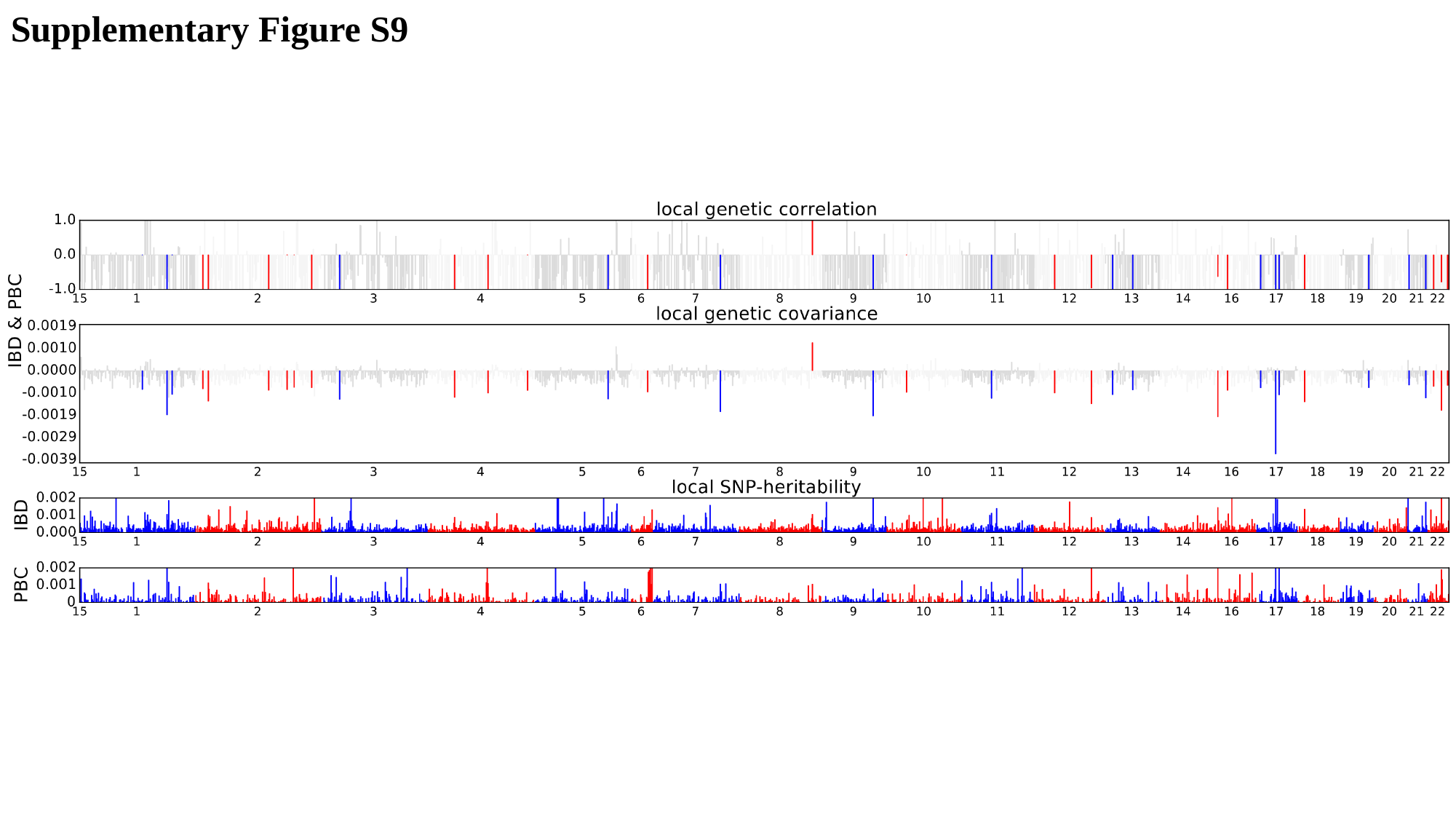

Supplementary Figure S9

### Slide 10
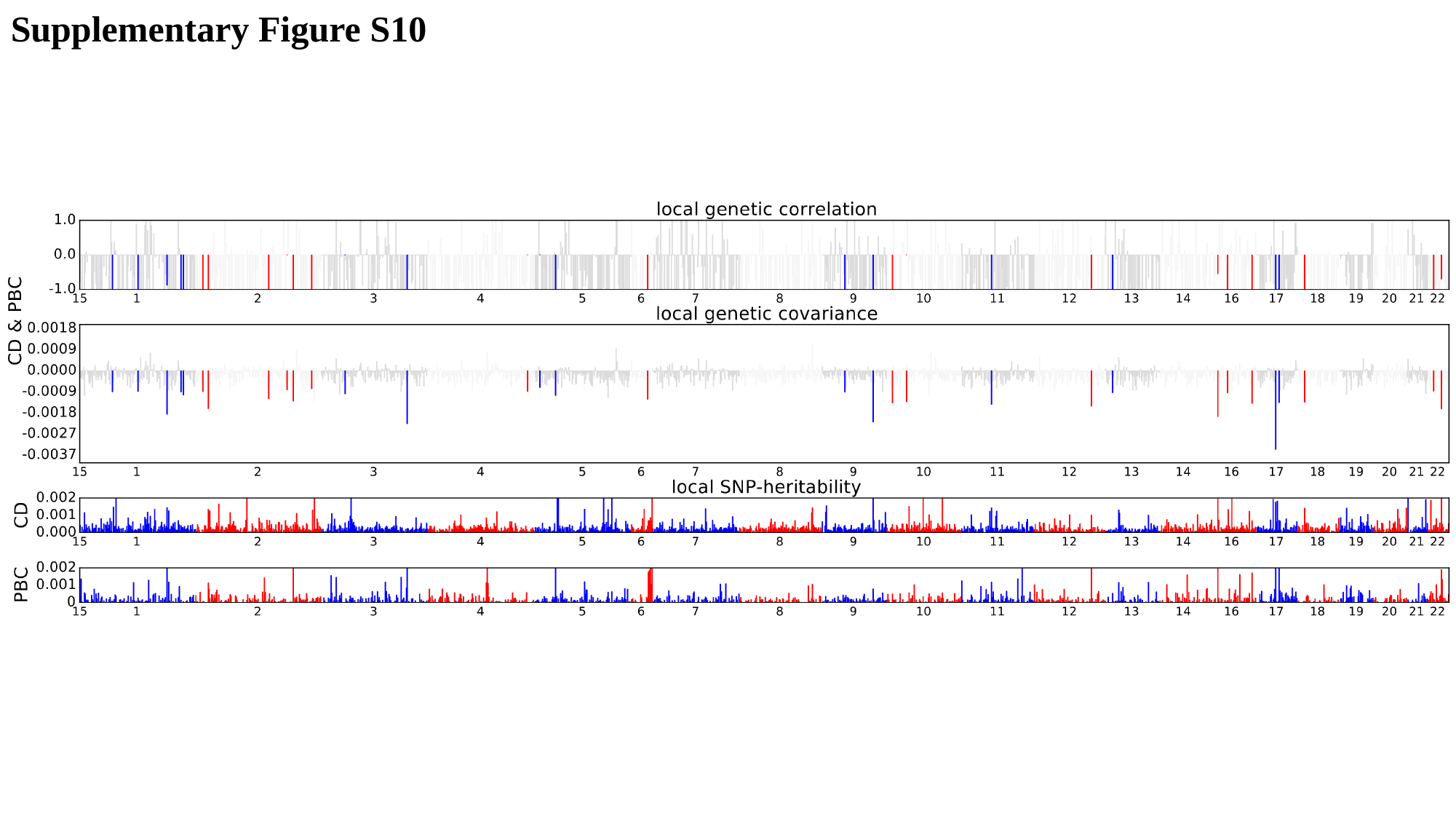

Supplementary Figure S10

### Slide 11
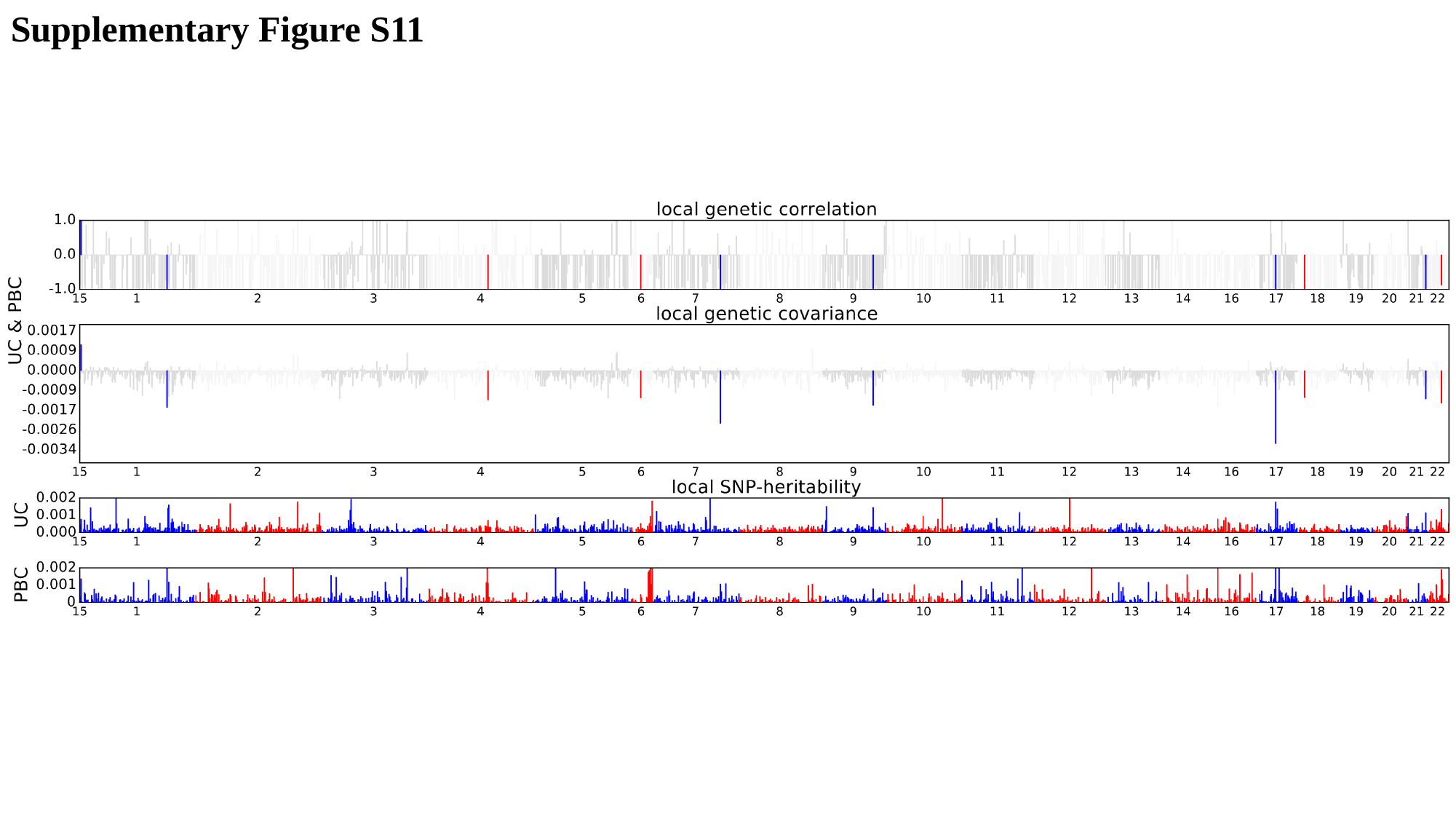

Supplementary Figure S11

### Slide 12
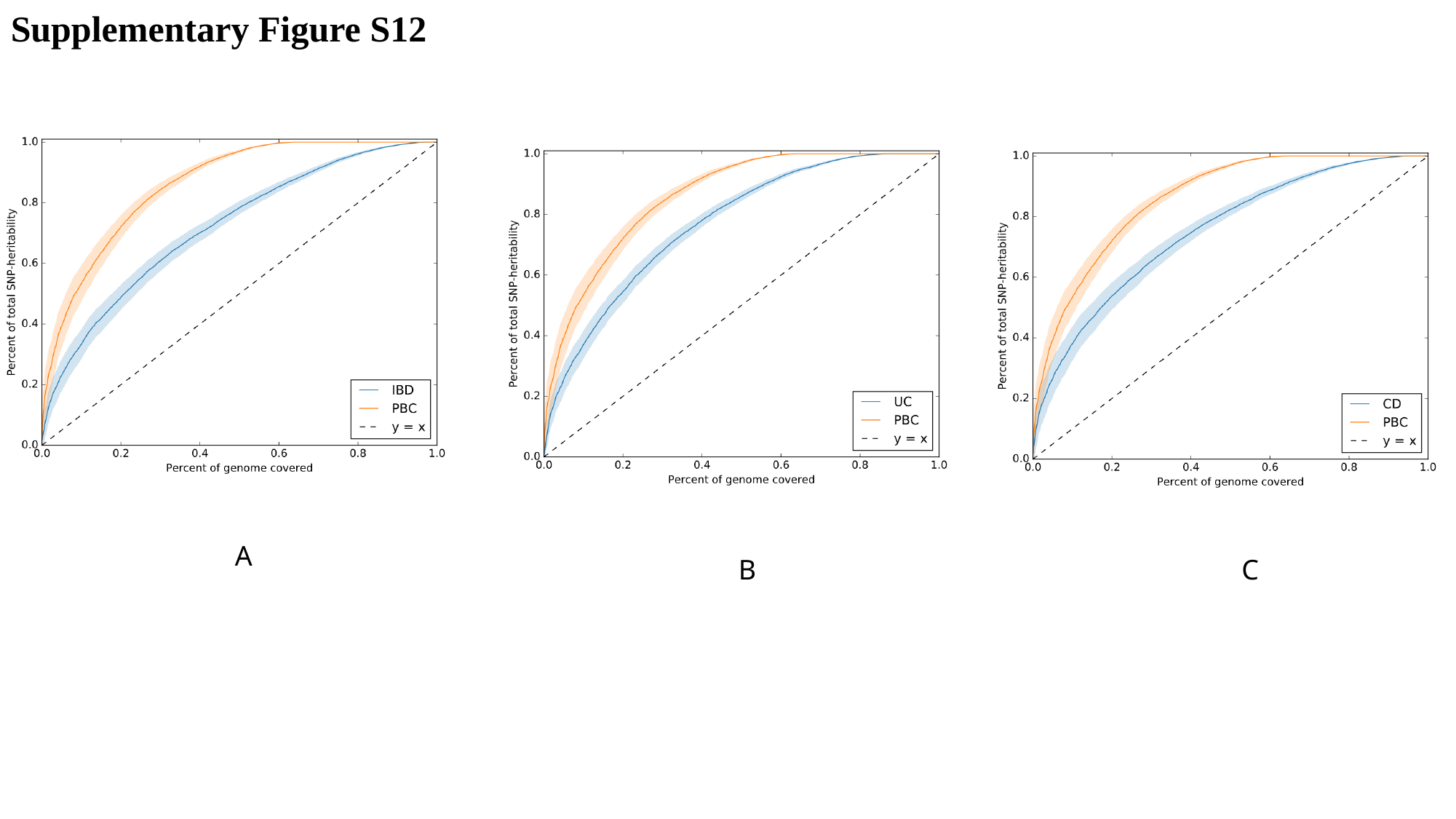

Supplementary Figure S12
A
B
C

### Slide 13
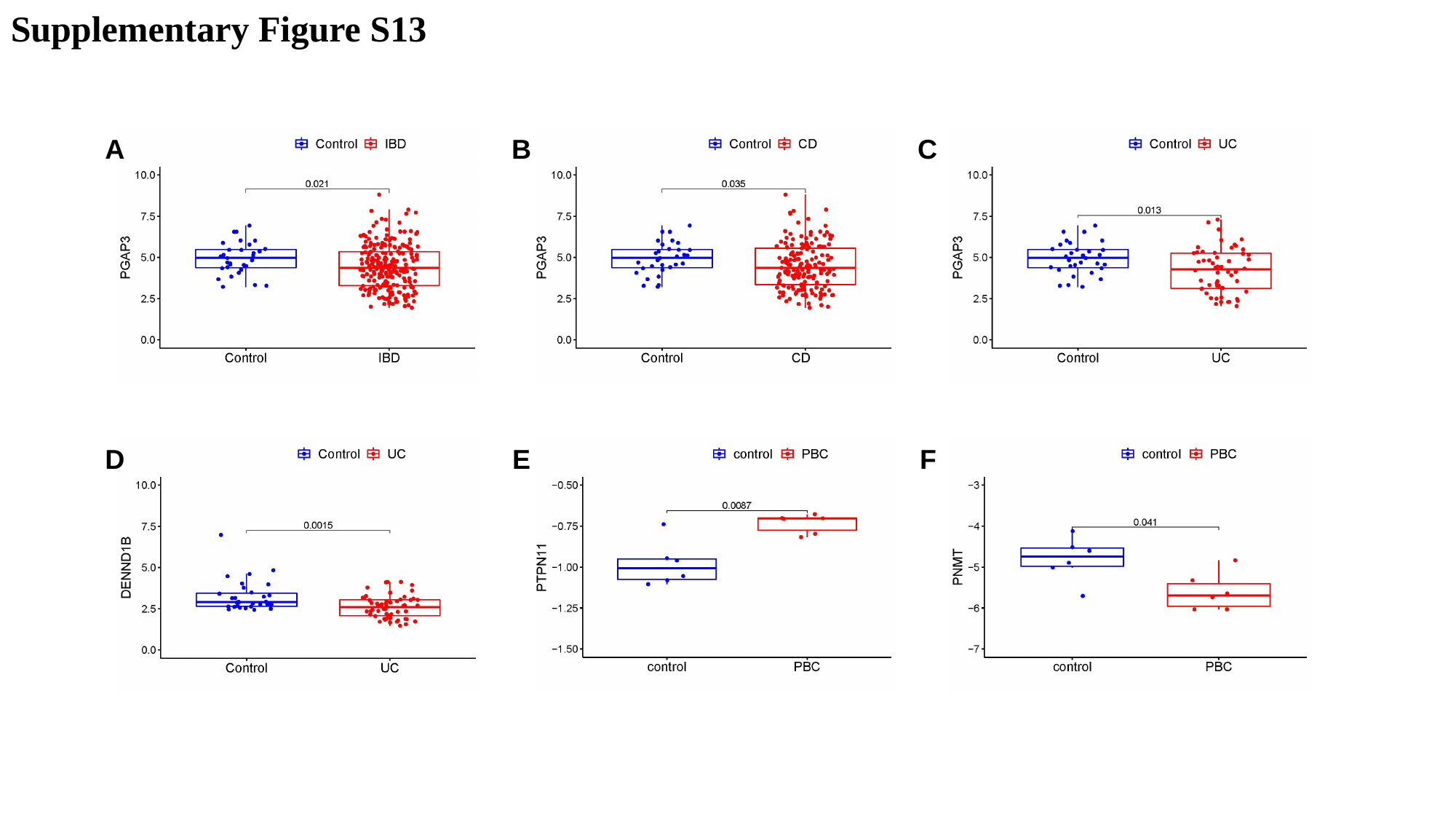

Supplementary Figure S13
A
B
C
D
E
F

### Slide 14
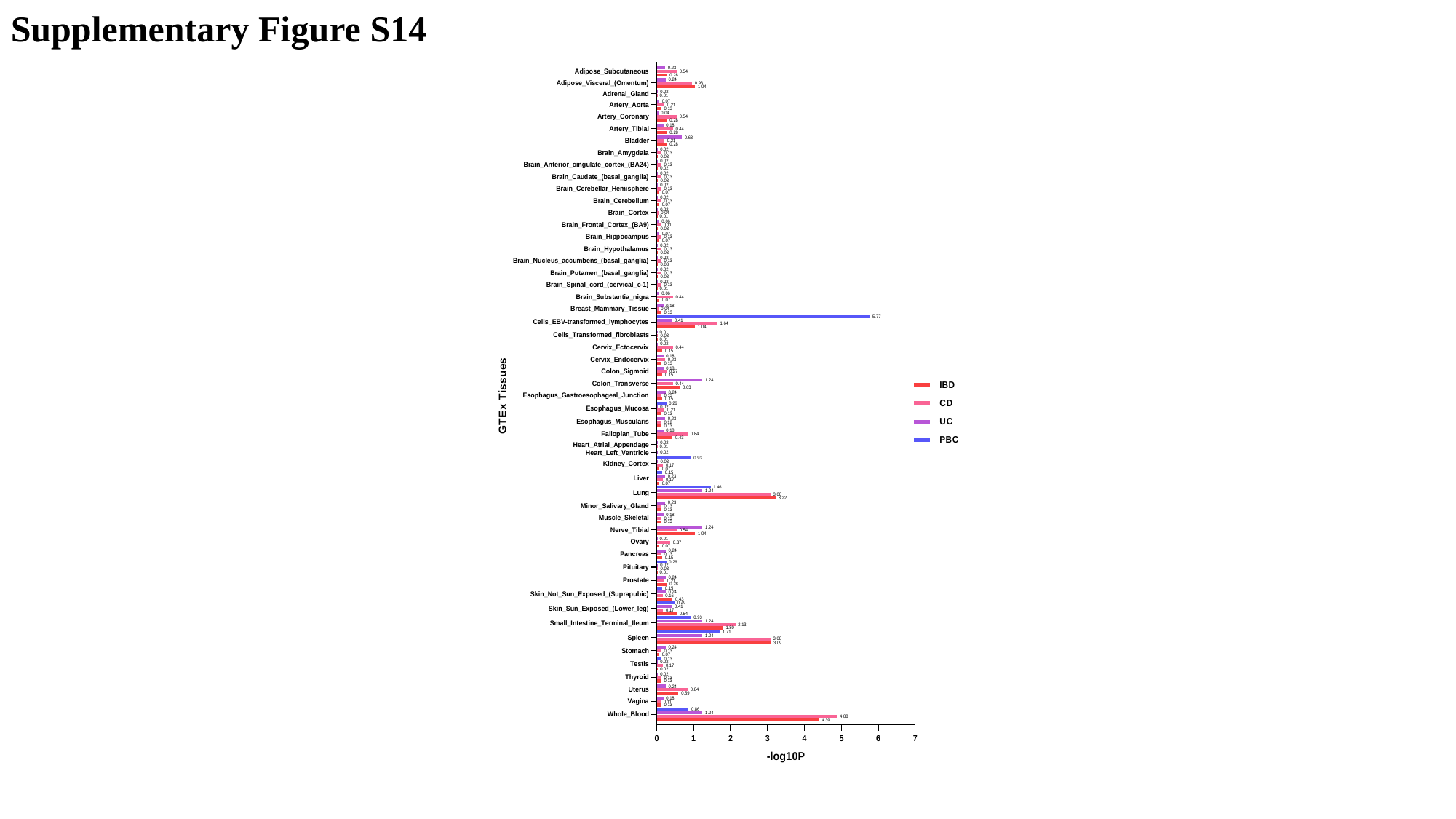

Supplementary Figure S14

### Slide 15
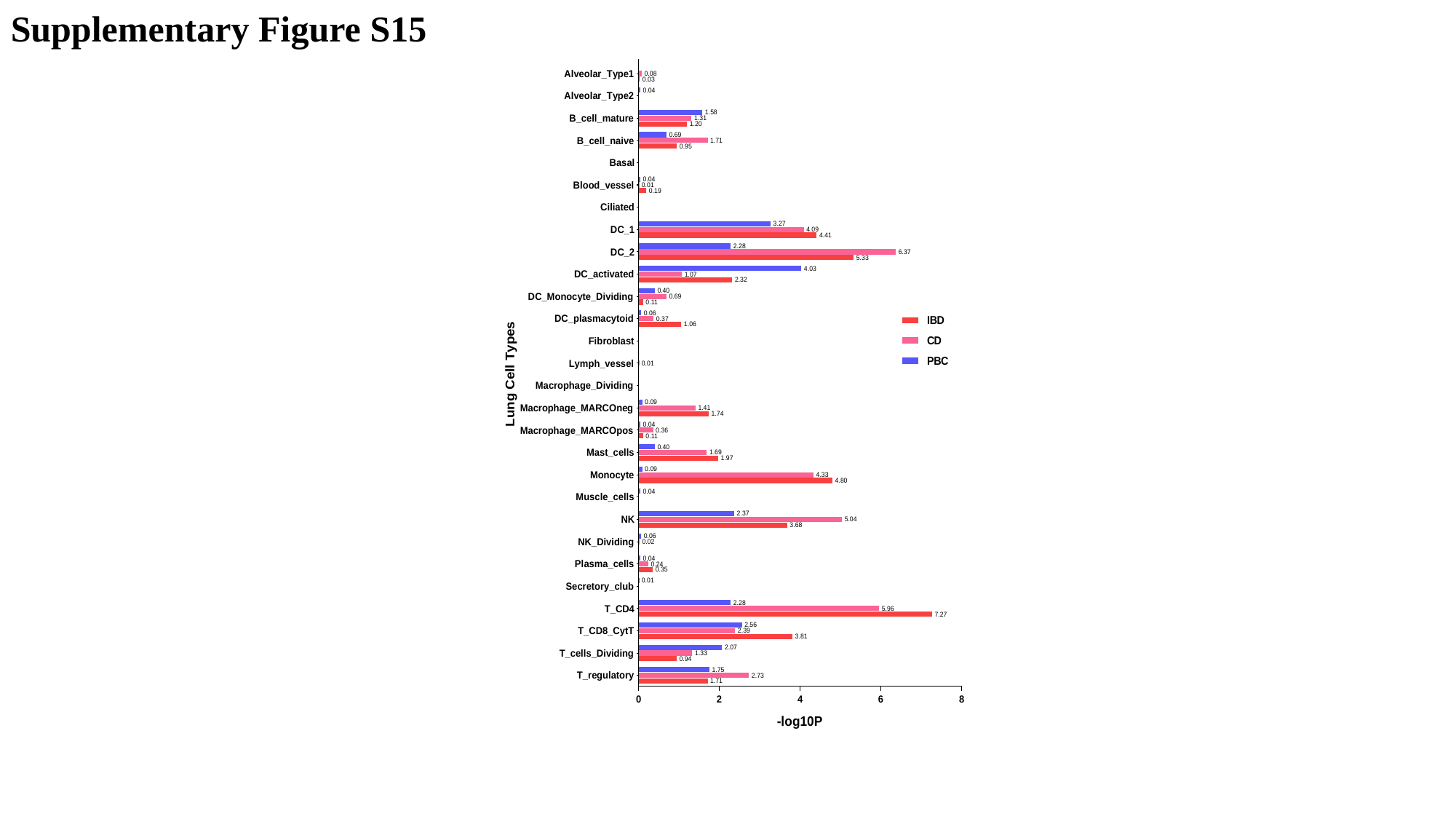

Supplementary Figure S15

### Slide 16
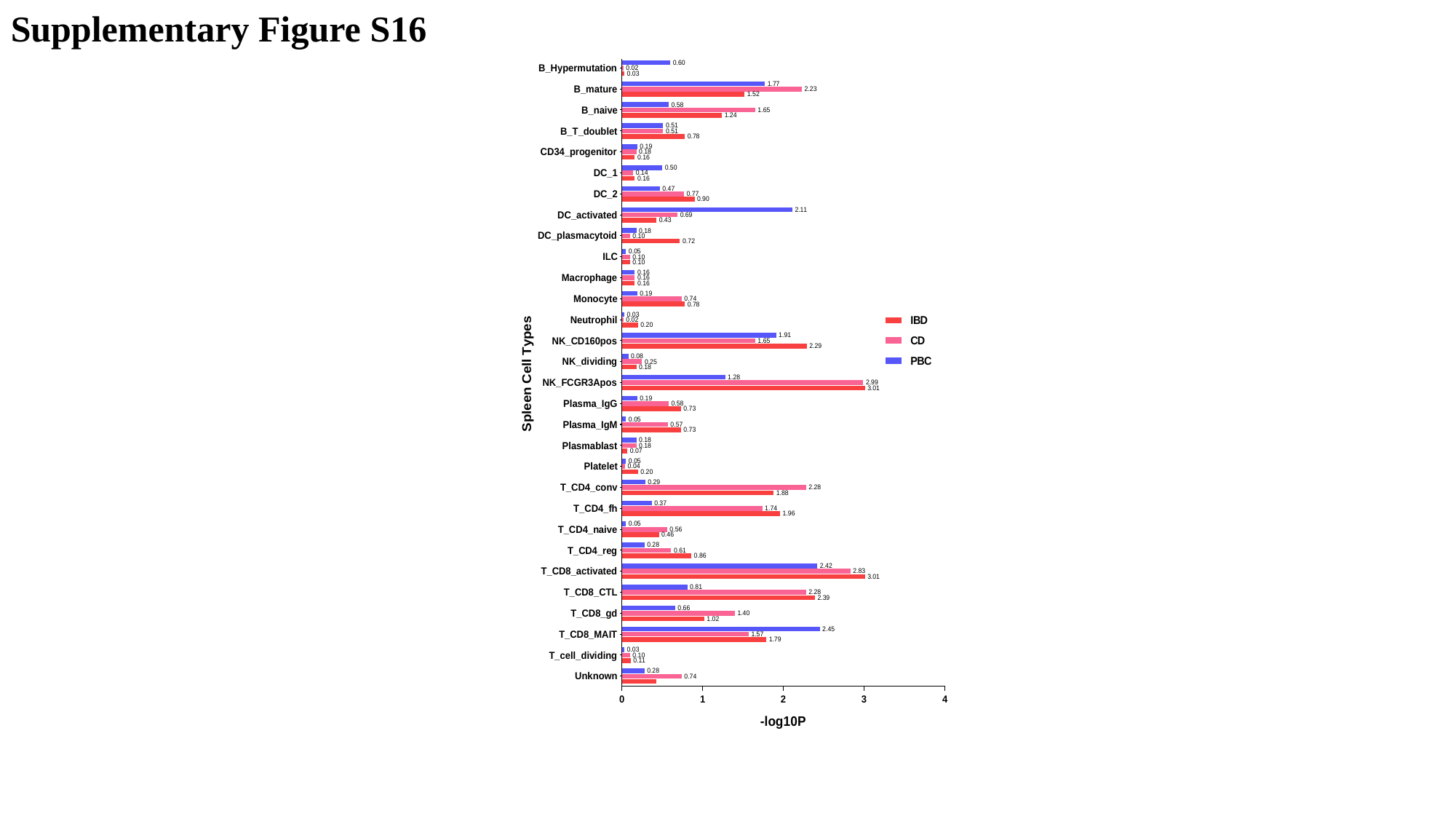

Supplementary Figure S16

### Slide 17
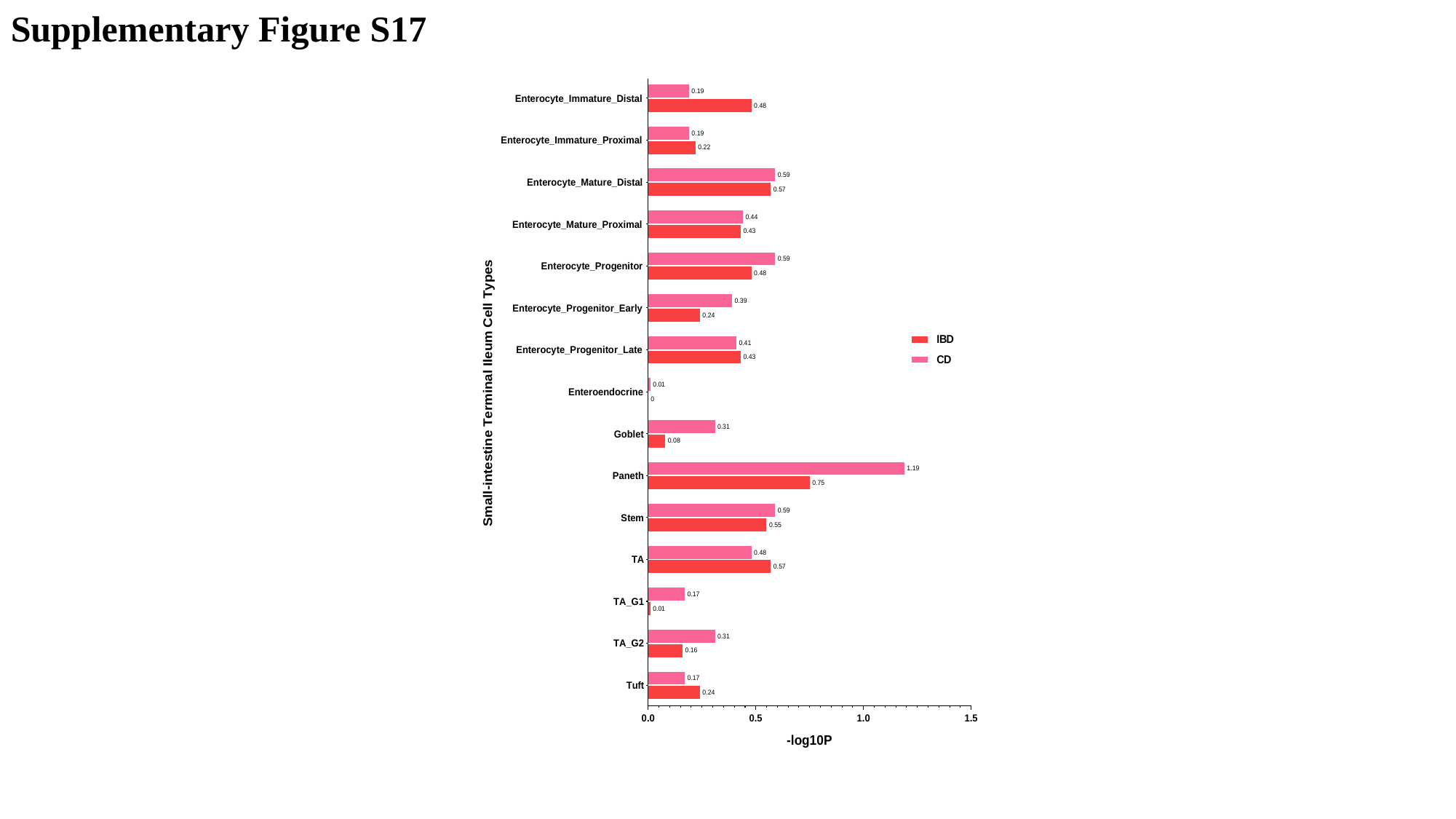

Supplementary Figure S17

### Slide 18
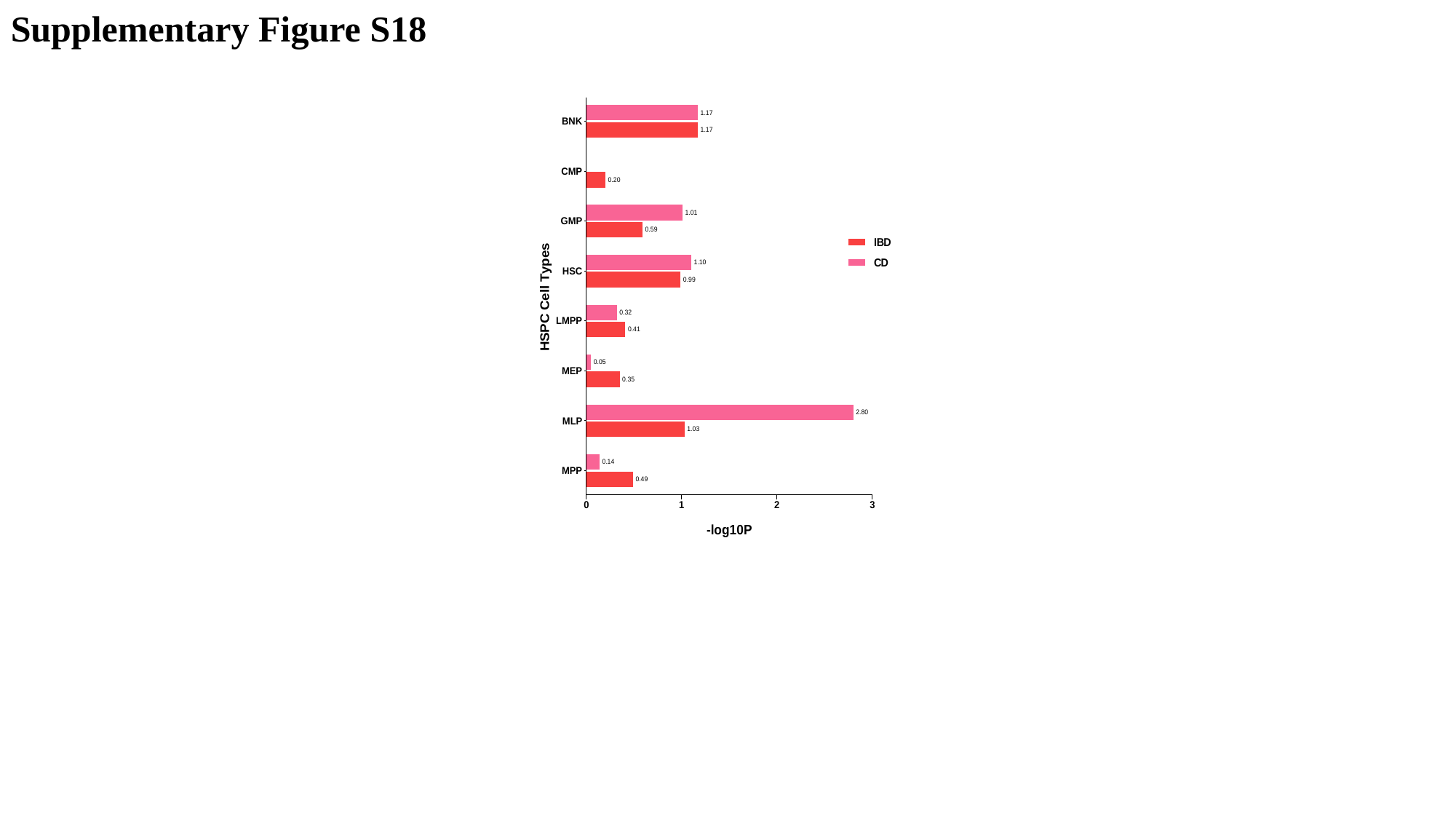

Supplementary Figure S18
